# Plasma multi-omic and cardiac imaging network signatures predict poor long-term outcomes after acute myocardial infarction

**DOI:** 10.1101/2022.04.08.22273590

**Authors:** Hiromi W.L. Koh, Anna Pilbrow, Sock Hwee Tan, Qing Zhao, Peter I. Benke, Bo Burla, Federico Torta, John W. Pickering, Richard Troughton, Christopher Pemberton, Wern-Miin Soo, Lieng Hsi Ling, Robert N. Doughty, Hyungwon Choi, Markus R. Wenk, A. Mark Richards, Mark Y. Chan

## Abstract

**Background:** Prognostic biomarkers for patients admitted for a myocardial infarction (MI) episode are of great interest for risk stratification and follow-up care after discharge. Multi-omics analysis is a standard approach for the discovery of diagnostic and prognostic biomarkers, but few studies have evaluated the prognostic potential of molecular markers in combination with echocardiographic imaging variables.

**Methods:** We measured the plasma proteome and lipidome in patients discharged from an acute MI and followed for secondary outcomes in New Zealand for a median time of 4.85 years (CDCS, N=741 for network inference, N=464 for predictive analysis) and in Singapore for a median time of 2.0 years (IMMACULATE, N=190 for validation). Using a network-based integrative analysis framework iOmicsPASS+, we mapped proteins, lipids, echocardiographic imaging variables and clinical biomarkers to a unified network and identified predictive subnetwork signatures of major adverse cardiac events (MACE) and heart failure hospitalization (HFH) in CDCS, with validation in IMMACULATE.

**Results:** Specific plasma proteins and lipids showed direct connections to cardiac imaging variables in the network. The gold standard biomarker, NT-proBNP, remained one of the best prognostic marker of MACE and HFH, but a number of plasma proteins involved in extracellular matrix organization, chemotaxis, inflammation, and apoptosis were also strong predictors of both outcomes. Hub proteins of subnetwork signatures were enriched in the heart, arteries, kidneys, liver and lungs. *BMP10, CAPG, EFEMP1, FSTL3, RSPO4*, and *RELT* were those directly connected to the echocardiographic variables and natriuretic peptides. In particular, *EFEMP1* and *FSTL3* in combination with diastolic function (E/e’) were strongly predictive of HFH in both CDCS (AUC 0.78, 95%CI 0.72-0.83) and IMMACULATE (AUC 0.72, 0.61-0.84).

**Conclusions:** Our integrative analysis revealed competing signatures beyond established biomarkers of post-MI HFH, comprised of plasma proteins correlated with impaired diastolic function after the primary MI episode.

## Introduction

Circulating proteins and peptides of cardiac and non-cardiac origin are clinically important biomarkers for diagnosis and prognosis in cardiovascular disease (CVD).^1-3^ Historically, clinical biomarker studies have favoured approaches focusing on one or a few prioritised candidates within a risk stratification framework for primary and secondary outcomes^4-7^. Recent trends in CVD biomarker studies have pivoted towards a more unbiased approach, simultaneously analysing hundreds or thousands of molecules, enabled by medium to high-throughput omics technologies such as mass spectrometry and affinity proteomics.^8-11^

Although novel assay platforms allow for data generation for different molecular types and the diversity propels the discovery of novel biomarker candidates, the large volume of data, heterogeneity of molecular variations across data types, and differential coverage of assay platforms altogether render the process of data interpretation a challenging task. The conventional approach to biomarker discovery is driven by metrics of classification accuracy such as sensitivity, specificity and area under the curve (AUC) of the receiver-operating characteristic (ROC). While this dogmatic practice is necessary for careful selection of biomarkers, some candidates with equally good diagnostic or prognostic values, even with direct biological connections to altered cardiac function and vasculature remodelling, may be neglected unless they outperform benchmark biomarkers in terms of the metrics.

For unbiased assessment of the proteome as a biomarker pool in the population of post-myocardial infarction (MI) patients, we have recently reported plasma proteomics data collected from post-MI patients 30 days after hospital discharge from two distinct populations: (1) the Coronary Disease Cohort Study (CDCS) in New Zealand and (2) the Improving Outcomes in Myocardial Infarction through Reversal of Cardiac Remodelling (IMMACULATE) registry in Singapore.^12^ This work demonstrated a promising workflow for prioritising plasma proteins predictive of post-MI heart failure (HF) based on a comparative analysis between the plasma proteomic data and a single cell-resolution transcriptomics data set of mouse heart tissues.

Expanding on this work, we have generated targeted mass spectrometry-based lipidomics data and curated cardiac imaging data 30 days after hospital discharge for the same patients in both cohorts. Together, the comprehensive data set consists of 1690 data features, including two omics-scale measurements of circulating lipids/acylcarnitines and proteins, four natriuretic peptides (ANP, BNP, NT-proANP, NT-proBNP), cardiac troponin I (hsTNI), creatinine, and 19 echocardiographic imaging variables acquired by standardised protocol. The data set therefore represents a comprehensive data resource to study the correlation structures among different data types (inter-modality correlations) and within each data type (intra-modality correlations), and evaluate their joint prognostic potential for secondary major adverse cardiovascular events (MACE) and, more specifically, heart failure hospitalization (HFH).

Given the highly correlated nature of data features within each modality, we tackled the multi-modal data integration problem via a network-based approach, termed iOmicsPASS+, which we implemented as an open-source R package. The workflow first identifies a network of conditionally dependent data features via estimation of a sparse precision matrix,^13^ and teases out predictive subnetwork signatures of clinical outcomes using a network-level scoring approach (iOmicsPASS).^14^ Throughout this work, we leveraged on this two-stage workflow as the main computational engine for data integration.

## Results

### Characteristics of the CDCS cohort

**Figure 1A** illustrates the analysis workflow for the CDCS cohort. Using iOmicsPASS+, we integrated data for proteins, lipids, clinical biomarkers and echocardiographic variables from 741 subjects by deriving a network of conditionally dependent data features using the graphical LASSO.^15^ To refer to the variables of diverse data types, we call them *data features* hereafter. For this network, we calculated partial correlation for each edge as a measure of association strength between the two corresponding data features, accounting for all other variables (see **Supplementary Information**). In the second step, we identified predictive subnetwork signatures of secondary adverse outcomes, namely MACE and HFH. This supervised analysis was conducted excluding 286 patients (**Supplementary Information**). Of those 464 patients,185 patients remained event-free, and 279 patients had a secondary MACE, including 117 patients hospitalised for HF during follow-up.

**Figure 1.**
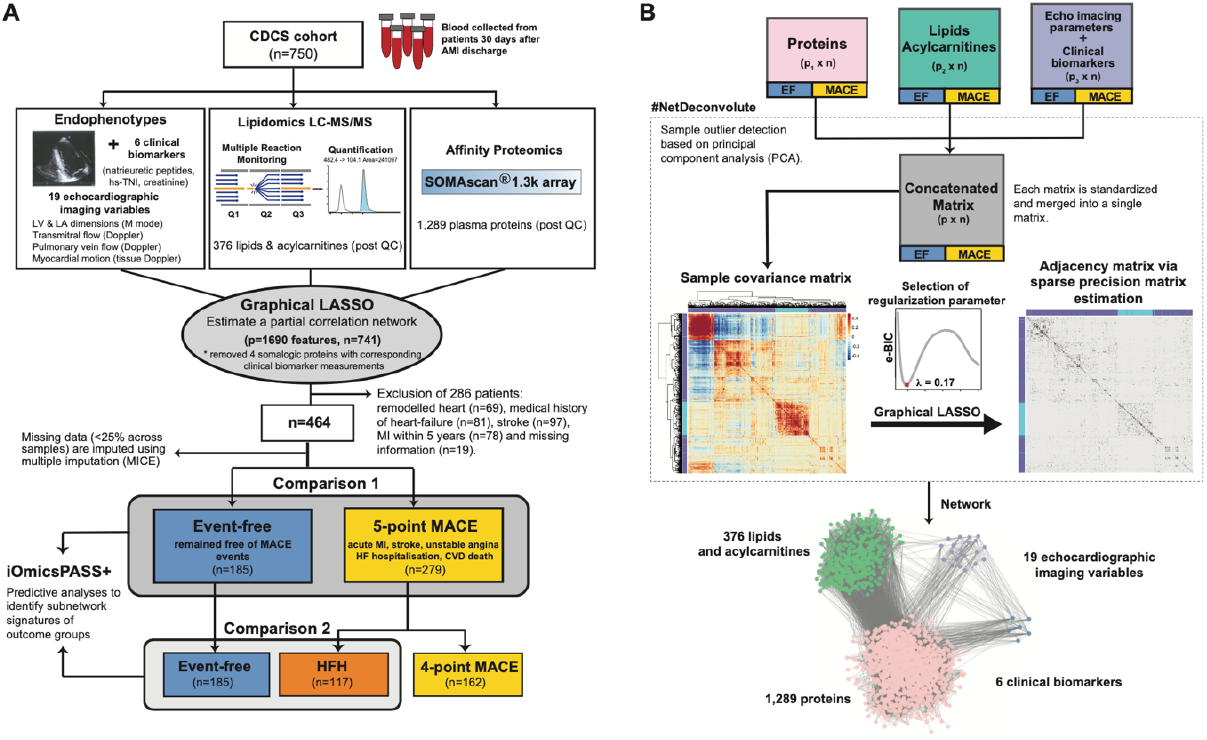
Analysis workflow applied to the CDCS cohort. (**A**) The background network reflecting the conditional dependence structure among the four data types is inferred using graphical LASSO. From the network, subnetwork signatures of secondary MACE and HFH are obtained using the training data (CDCS). (**B**) Data features are standardised and outliers are removed prior to integration of multi-modal data sets. In CDCS, 1,289 proteins from SOMALOGIC and 376 lipids from targeted MS were integrated with 19 cardiac imaging variables and 6 clinical biomarkers through this network inference.

**Table 1** provides the overall characteristics of the 464 patients. The mean age was 69 years (SD=10.7 years), with more males (69.2%) than females (30.8%). The majority were of European descent from New Zealand (56.7%) and other countries (27.8%), while the rest (7.1%) were Asians, Africans, Maoris, Fijians and the Pacific Islanders. Most patients either quit smoking (54.5%) or had never smoked (38.8%). Upon admission for a primary acute MI episode, 29.7 % were diagnosed with STEMI and 70.3% NSTEMI. The median follow-up time from hospital discharge was 4.88 years, and the time from discharge to a MACE ranged from one day to 8.8 years (median 0.74 years). Comparison between event-free patients and MACE patients revealed significant differences in age, diagnosis of ST-segment elevation and hypertension, as well as clinical biomarkers such as serum creatinine and plasma natriuretic peptides. MACE patients were older (mean age= 70.6 years, SD=11.1) and more frequently hypertensive (56.6%) than event-free patients (36.8%). The index events were also more likely NSTEMI (74.6%) than STEMI (63.8%). Event-free patients had lower levels of serum creatinine and plasma concentrations of natriuretic peptides (ANP, BNP, NT-proANP, NT-proBNP) than MACE patients on follow-up.

**Table 1.** Clinical characteristics of post-myocardial infarction patients in CDCS study

|  | CDCS study (New Zealand) |  |  | P-value |
| --- | --- | --- | --- | --- |
|  | All<br>(n = 464) | Event-free<br>(n = 185) | MACE<br>(n = 279) |  |
| **Follow-up time, median (yrs)** | 4.88 | 4.84 | 4.96 |  |
| **Time to MACE, median (yrs)** |  | - | 0.74 |  |
| **Age (yrs), mean (SD)** | 69 (10.7) | 66.7 (9.6) | 70.6 (11.1) | <0.001 |
| **BMI (kg/m<sup>2</sup>), mean (SD)** | 27.1 (4.9) | 27 (3.9) | 27.2 (5.5) | 0.603 |
| **Gender, n (%)** |  |  |  |  |
| Males | 321 (69.2) | 126 (68.1) | 195 (69.9) | 0.760 |
| Females | 143 (30.8) | 59 (31.9) | 84 (30.1) |  |
| **Ethnicity, n (%)** |  |  |  |  |
| NZ European | 263 (56.7) | 102 (55.1) | 161 (57.7) | 0.485 |
| Other Europeans | 129 (27.8) | 42 (22.7) | 87 (31.2) |  |
| Others | 33 (7.1) | 12 (6.5) | 21 (7.5) |  |
| Unknown | 39 (8.4) | 29 (15.7) | 10 (3.6) |  |
| **Smoking status, n (%)** |  |  |  |  |
| Current Smoker | 31 (6.7) | 13 (7) | 18 (6.5) | 0.318 |
| Ex-Smoker | 253 (54.5) | 93 (50.3) | 160 (57.3) |  |
| Never Smoked | 180 (38.8) | 79 (42.7) | 101 (36.2) |  |
| **Alcohol consumption, n (%)** |  |  |  |  |
| Current drinker | 294 (63.4) | 127 (68.6) | 167 (59.9) | 0.070 |
| Ex-drinker | 52 (11.2) | 14 (7.6) | 38 (13.6) |  |
| non-drinker | 118 (25.4) | 44 (23.8) | 74 (26.5) |  |
| **ST-elevation status, n (%)** |  |  |  |  |
| ST-elevated MI, STEMI | 138 (29.7) | 67 (36.2) | 71 (25.4) | 0.017 |
| Non ST-elevated MI, NSTEMI | 326 (70.3) | 118 (63.8) | 208 (74.6) |  |
| **Family history of CAD, n (%)** | 185 (39.9) | 80 (43.2) | 105 (37.6) | 0.345 |
| **Diabetes Mellitus, n (%)** | 82 (17.7) | 27 (14.6) | 55 (19.7) | 0.197 |
| **Hypertension, n (%)** | 226 (48.7) | 68 (36.8) | 158 (56.6) | <0.001 |
| **Hyperlipidemia, n (%)** | 209 (45) | 82 (44.3) | 127 (45.5) | 0.687 |
| **Clinical- biomarkers<sup>‡</sup>, mean (SD)** |  |  |  |  |
| Creatinine, mg/dL | 99.4 (59.7) | 90.3 (18.7) | 106 (75.2) | <0.001 |
| high-sensitive Troponin I (hsTNI), ng/L | 57.8 (506.6) | 18.3 (51.5) | 84.6 (654.0) | <0.001 |
| Atrial natriuretic peptide (ANP), pg/mL | 401 (271.8) | 347 (210.5) | 436 (301) | 0.001 |
| N-terminal pro ANP, pg/mL | 12200 (8652.3) | 9830 (5839.7) | 13700 (9810.5) | <0.001 |
| Brain natriuretic peptide (BNP), pg/mL | 260 (283.5) | 187 (198.8) | 310 (319.0) | <0.001 |
| N-terminal pro BNP, pg/mL | 1330 (1539.8) | 925 (908.6) | 1600 (1795.6) | <0.001 |

\* P-values of the comparisons across the three groups (Event-free, MACE and HFH patients) where ANOVA is used for continuous variables and chi-square test is used for categorical variables.
† Markers were measured during hospital admission for a primary MI event.
‡ Markers were measured one month from hospital discharge post MI.
CDCS, Coronary artery Disease Cohort Study; CAD, coronary artery disease; NZ, New Zealand; BMI, body mass index; MI, myocardial infarction; HF, heart failure; LVEF, left ventricular ejection fraction; STEMI, ST-elevated myocardial infarction; NSTEMI, non ST-elevated myocardial infarction; LDL, low-density lipoprotein; HDL, high-density lipoprotein; hsTNI, high-sensitive Troponin I; NT-proANP, N-terminal pro-hormone atrial natriuretic peptide; NT-proBNP, N-terminal pro-hormone brain natriuretic peptide.

**Table 2** shows the comparison of the echocardiographic variables between event-free patients and MACE patients, as well as between event-free patients and HFH patients. As expected, the echocardiographic variables differed more between HFH patients and event-free patients than between all MACE patients and even-free patients. Both adverse outcome groups (MACE and the HFH subgroup) had greater baseline LV dimensions and lower left ventricle ejection fraction (LVEF) than event-free patients. Tissue Doppler variables of myocardial motion differed between MACE/HFH patients and event-free patients with e’ and E/e’ differing most between the HFH subgroup and event-free patients.

**Table 2.** Echocardiographic imaging variables of post-myocardial infarction patients in CDCS study

| CDCS study | Event-free,<br>EF<br>(n = 185) | MACE<br>(n = 279) | P-value for<br>EF vs<br>MACE | HFH<br>(n = 117) | P-value for<br>EF vs HFH |
| --- | --- | --- | --- | --- | --- |
| **Heart dimensions** |  |  |  |  |  |
| Interventricular septum, IVS (mm) | 11.4 (2.2) | 12.1 (2.9) | 0.008 | 12.4 (2.8) | 0.001 |
| Posterior wall thickness, PWT (mm) | 11 (1.9) | 11.5 (2.9) | 0.105 | 11.5 (2.3) | 0.106 |
| Left ventricle (LV) mass, LVMI (g/m<sup>2</sup>) | 124 (31.1) | 141 (46.6) | <0.001 | 148 (39.0) | <0.001 |
| Left ventricle internal dimension diastole, LVIDDi (mm/m<sup>2</sup>) | 28.1 (3.8) | 29.2 (4.3) | 0.008 | 30.2 (5.0) | <0.001 |
| Left ventricle internal dimension systole, LVIDSi (mm/m<sup>2</sup>) | 18.8 (3.9) | 19.9 (4.6) | 0.010 | 21.4 (5.0) | <0.001 |
| Left ventricle end diastolic volume, LVEDVi (mL/m<sup>2</sup>) | 64.4 (15.3) | 66.9 (20.9) | 0.211 | 69.0 (22.4) | 0.060 |
| Left ventricle end systolic volume, LVESVi (mL/m<sup>2</sup>) | 27.7 (12.1) | 31.6 (17.6) | 0.021 | 35.7 (20.3) | <0.001 |
| Left ventricle ejection fraction, LVEF (%) | 58.4 (10.8) | 56 (12.4) | 0.032 | 51.7 (13.6) | <0.001 |
| Left Atrial (LA) width (mm) | 40.3 (5.9) | 41.8 (7) | 0.018 | 44.3 (6.9) | <0.001 |
| Left Atrial (LA) area (cm<sup>2</sup>) | 20.7 (5.7) | 23 (5.6) | <0.001 | 24.6 (6.2) | <0.001 |
| **Transmitral flow** |  |  |  |  |  |
| Deceleration Time, DT (msec) | 238 (67.3) | 237 (73.8) | 0.865 | 229 (77.5) | 0.288 |
| Peak E/A ratio | 1.11 (0.6) | 1.04 (0.4) | 0.195 | 1.05 (0.5) | 0.421 |
| **Pulmonary vein flow** |  |  |  |  |  |
| Peak S/D ratio | 1.39 (0.5) | 1.32 (0.4) | 0.145 | 1.23 (0.5) | 0.017 |
| Peak AR (cm/s) | 28.7 (7.8) | 28.1 (7.7) | 0.415 | 28.8 (8.4) | 0.942 |
| **Tissue Doppler imaging** |  |  |  |  |  |
| e' (cm/s), average of lateral and septal annuli | 8.1 (2.2) | 7.6 (2.2) | 0.027 | 7.2 (2.3) | 0.001 |
| a' (cm/s), average of lateral and septal annuli | 9.3 (2.1) | 9.1 (2.4) | 0.368 | 8.6 (2.3) | 0.005 |
| s' (cm/s), average of lateral and septal annuli | 7.6 (1.4) | 7.4 (1.7) | 0.288 | 6.9 (1.6) | <0.001 |
| e'/a' ratio | 0.91 (0.3) | 0.89 (0.4) | 0.519 | 0.88 (0.4) | 0.485 |
| E/e' ratio | 9.2 (3.1) | 10.7 (4.2) | <0.001 | 12.2 (5.0) | <0.001 |

### Single markers of secondary MACE and HFH

We next compared the levels of individual proteins, lipids, echocardiographic variables and clinical biomarkers in the patients with MACE and HFH to event-free patients. A total of 184 data features were significantly different in mean (FDR<0.05) between MACE patients and event-free patients, where 66.3% were higher in MACE patients. Comparing the HFH patients with the event-free ones, 368 markers were significantly different (FDR<0.05). Details of this analysis are reported in **Supplementary Table 1**.

The differential features of MACE included 166 proteins, seven lipids, five echocardiographic variables, and all six clinical biomarkers. Although all natriuretic peptides, cardiac troponin I (hsTNI) and creatinine levels were higher in MACE patients, BNP and NT-proBNP attained the AUC of 0.65 (95% CI: 0.60-0.70 for BNP and 0.59 – 0.70 for NT-proBNP), indicating modest discrimination for MACE. Echocardiographic variables such as left atrial (LA) area, left ventricle (LV) mass, indexed left ventricle internal dimension in diastole (LVIDDi) and systole (LVIDSi), as well as indices of diastolic dysfunction (E/e’), were all higher in MACE patients than in event-free ones, yet the AUC values were modest at best. When considering lipids, three (phosphatidylethanolamines PE 34:1 and PE 34:2, sphingosine-1-phosphate S1P d18:0) were higher, whereas four species (phosphatidylcholine PC 38:4 and 40:8, lysophosphatidylcholine LPC 20:4, sphingomyelin SM 43:1) were lower in the MACE patients. Lastly, the top three markers were all plasma proteins, including macrophage-capping protein (*CAPG*) with AUC of 0.68 (95% CI: 0.63 – 0.73), aspartate aminotransferase (*GOT1*) with AUC of 0.65 (95% CI: 0.60 – 0.70) and follistatin-related protein 3 (*FSTL3*) with AUC of 0.64 (95% CI: 0.59 – 0.69).

The 368 differential features of HFH included 298 proteins, 31 lipids, nine echocardiographic variables, and all six clinical biomarkers. Similar to MACE, all six clinical biomarkers were higher in the HFH patients than in event-free ones as expected: NT-proBNP demonstrated the highest AUC of 0.79 (95% CI: 0.74 – 0.84), followed by BNP with AUC of 0.78 (95% CI: 0.73 – 0.83). In echocardiographic variables, all five variables significantly different in MACE also differed in the HFH subgroup. In addition, interventricular septum (IVS), LA width, LVESVi were also significantly higher, while the early diastolic mitral annulus velocity e’ was significantly lower. The significant lipids included glycerophospholipids (8 PEs, 3 PC, 4 phosphatidylinositol PI), four lysophospholipids (3 LPC, 1 LPE), eight sphingolipids (SM 38:1, 38:2, 43:1, 44:1, 44:2), S1P d18:0, ceramide d19:1/24:0 and ganglioside GM3 d18:1/16:0, cholesteryl ester CE 20:4 and two glycerolipids (diacylglycerol DG 38:6 and triacylglycerol TG 58:10). Among those, LPC 20:4, PE 34:2 and PE 35:2 had the highest AUCs, albeit at a modest value of 0.64 (95% CI: 0.57 – 0.71). Of the 298 significant proteins, 61.7% were higher in HFH patients than in event-free patients. *CAPG* had the highest AUC of 0.77 (95% CI: 0.72 – 0.83), followed by *FSTL3* with AUC of 0.75 (95% CI: 0.70 – 0.81) and Cystatin-C (*CST3*) with AUC of 0.74 (95% CI: 0.68 – 0.79). Together, although plasma proteins offered a slightly weaker classification power than the natriuretic peptides, best predictive proteins such as *CAPG, CST3, FSTL3*, and *EFEMP1* were equally predictive of HFH with overlapping 95% CIs.

### Network of data features from the four modalities

Next, we modified the integrative analysis approach called iOmicsPASS,^14^ originally developed for the integration of genomic, transcriptomic, and proteomic data over experimentally validated biological networks. The new implementation, iOmicsPASS+, is freely available to the public and amenable to general applications without pre-existing inter-molecular networks, such as studies of circulating biomarkers (see **Supplementary Information**). Compared to the previous implementation, the new interface offers greater flexibility to integrate diverse types of data and infer a network of conditional dependence among continuously scaled data features. iOmicsPASS+ can handle data sets with missing entries (up to 50%) and compute a sample covariance matrix using pairwise complete observations in order to retain as many data features and samples for downstream analysis. The network inference is enabled by sparse precision matrix estimation,^13^ which has a well-established foundation as Gaussian graphical models.^16^

**Figure 1B** illustrates the network inference workflow of iOmicsPASS+. First, each data type was standardised and outlier observations were filtered out before being concatenated into a single data matrix. The network estimation module produced a network of 27,334 edges in this data set (1.9% of all possible edges), connecting 1,690 data features. To assign measures of conditional dependence to the selected edges, the regularised estimates of precision values were converted to partial correlations, denoted by *r* (see **Supplementary Information**). The values ranged from -0.43 to 0.66 in this study. We remark that the reported partial correlation values are regularised estimates, i.e. not unbiased estimates.

Not surprisingly, the intra-modality correlations (within the same data type) were stronger than inter-modality correlations (between data types): only 5.4% of the edges connected proteins with lipids, and 373 edges connected proteins with echocardiographic variables and clinical biomarkers. This result shows that the proteomic variation is largely independent of the lipidomic variation and imaging variables, and both types of molecular data relate differently to the risk of future adverse outcomes.

The core segment of the network connecting echocardiographic variables and clinical biomarkers to proteins, lipids, and acylcarnitines is visualised in **Figure 2**, using the Cytoscape software.^17^ After accounting for the indirect associations explained away by other data features, natriuretic peptides and hsTNI were positively correlated with each other and only BNP, NT-proBNP and hsTNI were negatively correlated with LVEF. Among the echocardiographic variables, LVEDVi and LVESVi had the highest correlation (*r* = 0.475), followed by LVIDSi with LVIDDi (*r* = 0.459) and the negative correlation between LVEF and LVESVi (*r* = -0.375). Overall, only 10 echocardiographic variables were connected with clinical biomarkers in this subnetwork. The highest correlations were recorded between NT-proBNP and E/e’ (*r* = 0.058), between NT-proBNP and s’ (*r* = -0.057), between BNP and LVEF (*r* = -0.053) and between BNP and a’ (*r* = -0.050). This finding suggests that myocardial motion variables measured by tissue Doppler imaging are particularly correlated with neurohormonal activation from primary MI episodes.

**Figure 2.**
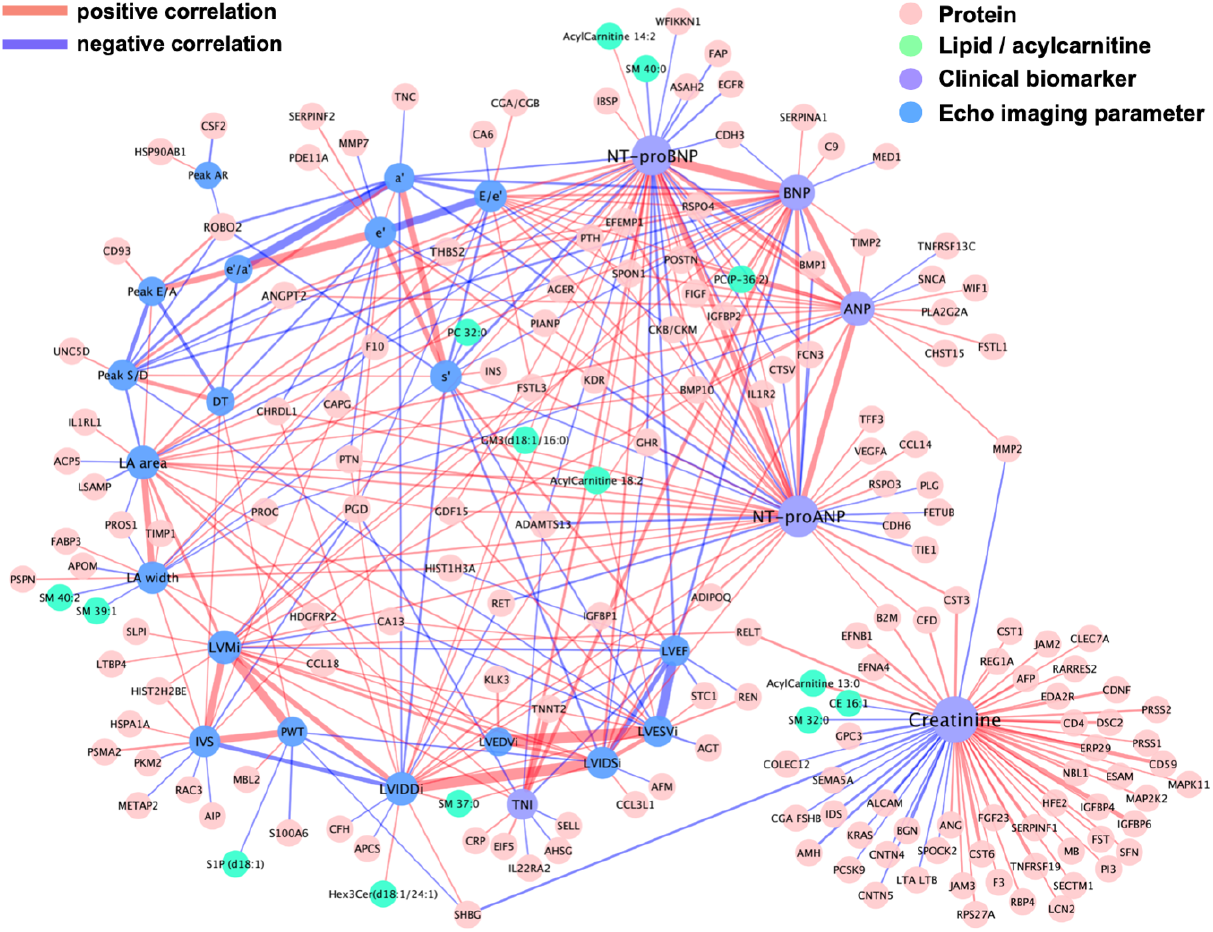
Visualization of the network connecting plasma proteins and lipids with echocardiographic imaging variables and cardiac biomarkers. Plasma proteins and lipids were drawn as nodes in light red and cyan, and echocardiographic variables and cardiac biomarkers in blue and light purple, respectively. Network edges were coloured according to the signs of partial correlations, i.e. positive in red and negative in light purple.

Meanwhile, stronger correlations were observed among proteins, lipids, and acylcarnitines with clinical biomarkers than with echocardiographic variables. In the lipidomics assay, the highest correlation was between acylcarnitine C13:0 and creatinine (*r* = 0.040). Two acylcarnitines (C14:2, C18:2) and two lipids (GM3 d18:1/16:0, PC-P-26:2) were positively correlated with NT-proBNP, whereas SM 40:0 was negatively correlated with NT-proBNP. PC-P 26:2 was the only lipid positively correlated with all four natriuretic peptides, PC 32:0 was negatively correlated with echocardiographic imaging variable s’, and acylcarnitine C18:2 was positively correlated with both LA area and LVESVi. Sphingomyelin (SM) 37:0 was positively correlated with internal diameters LVIDSi and LVIDDi, while SM 39:1 and SM 40:2 were negatively correlated with LA width.

Regarding the proteins, the highest correlation was between hsTNI and cardiac troponin T (*TNNT2*) (*r* = 0.206), followed by the correlation between creatinine and insulin-like growth factor binding protein 2 (*IGFBP2*) (*r* = 0.142), glycoprotein *CD59* (*r* = 0.118), and cystatin M (*CST6*) (*r* = 0.115). Nine proteins were correlated with three or more natriuretic peptides, including positive correlation with angiopoietin-2 (*ANGPT2*), periostin (*POSTN*), R-spondin 4 (*RSPO4*), insulin-like growth factor binding protein 2 (*IGFBP2*), thrombospondin 2 (*THBS2*), Spondin-1 (*SPON1*), vascular endothelial growth factor D (*VEGFD)* (also known as *FIGF*), bone morphogenetic protein 10 (*BMP10*), and negative correlations with bone morphogenetic protein 1 (*BMP1*) and coagulation factor X (*F10*). See **Supplementary Table 2** for the table of partial correlations among the data features from the four modalities.

### Subnetwork signature of MACE

Using this network as the background, we next applied the supervised analysis module of iOmicsPASS+ to obtain subnetwork signatures of MACE and HFH.^14^ **Table 2** reports the number of proteins, lipids, echocardiographic variables, and clinical biomarkers in the predictive signatures. The MACE signature included 524 edges connecting 211 nodes (cross-validated error 38.5%). **Figure 3** visualises the MACE signature, where the edges were coloured by the sign of the test-statistics (*d*_*ik*_^*^) for the MACE group (Koh *et al*^*14*^ for the details of the test statistics as group-specific centroids). The statistics, calculated by iOmicsPASS, are equivalent to group-specific centroids of the network, indexed for edge *i* for group *k* (*k*=1 for MACE, *k*=0 for event-free). The same network with the edges coloured by the signs of partial correlations between connected nodes is in **Supplementary Figure 1A**. This subnetwork signature contains 194 proteins, five lipids, eight echo imaging variables and all four natriuretic peptide markers. Although only five lipids were part of this network, four of them were phosphatidylethanolamines (PE 34:1, 34:2, 35:2, 37:4).

**Figure 3.**
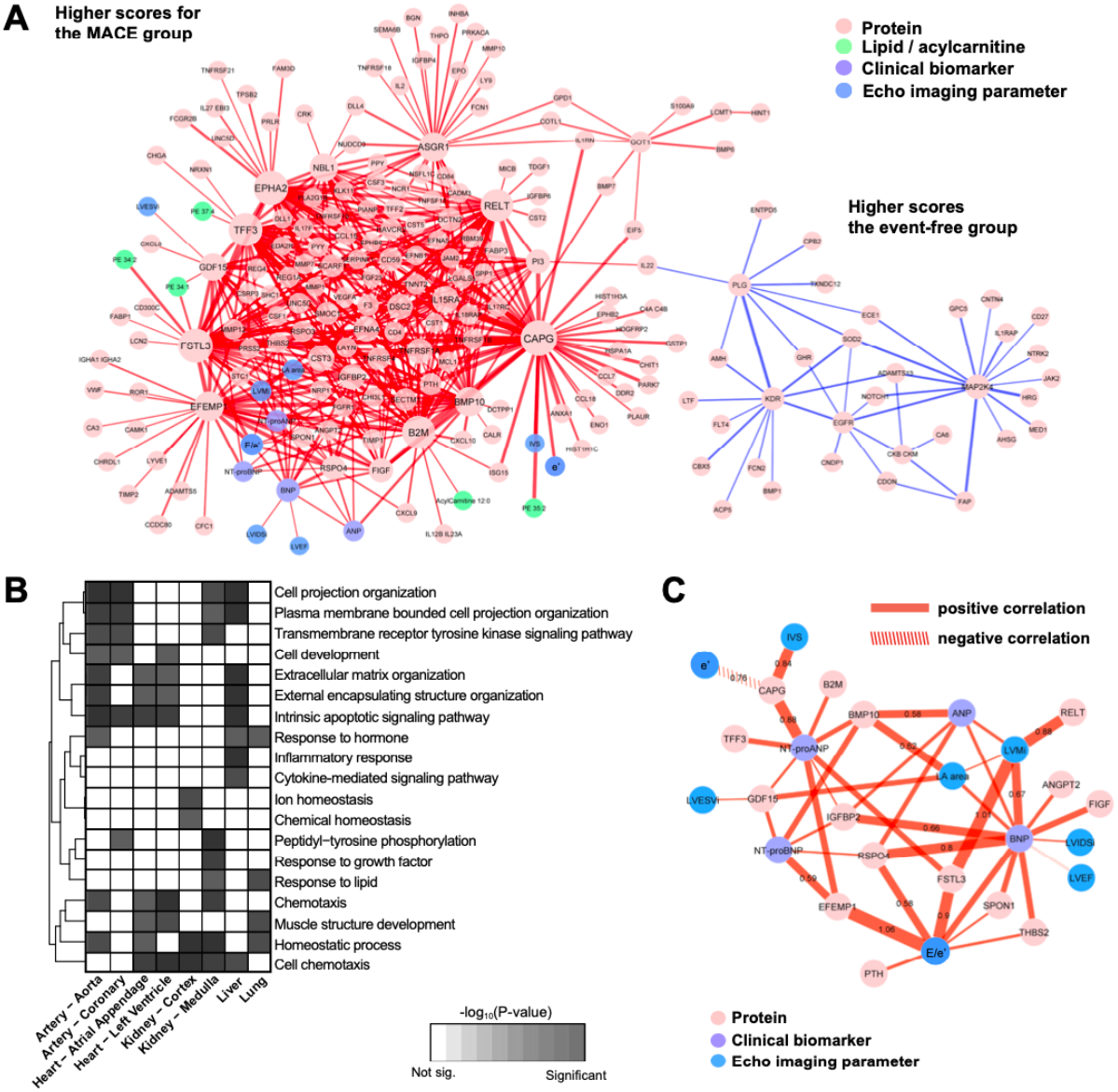
Subnetwork signatures of secondary MACE. (**A**) Network visualization with plasma proteins, lipids and acylcarnitines, echocardiographic imaging variables, and clinical biomarkers in different colours. Edges were coloured according to the signs of the group-specific test statistics (scaled centroids) computed by iOmicsPASS+. (**B**) Enrichment of biological functions in the constituent nodes in panel (**A**), accounting for tissue-specific expression of genes at the mRNA level (TPM > 5 in each tissue). (**C**) Visualization of the subnetwork connecting plasma proteins and echocardiographic imaging variables as well as clinical biomarkers.

The subnetwork was largely partitioned into two segments. The majority (96.4%) were highly correlated proteins, lipids, imaging variables and clinical biomarkers with higher d_ik_^*^ scores (in red) for the MACE group. The other part was a protein-only network with lower d_ik_^*^ scores (in blue). Here, the main driver of separation between the two groups were the edges connecting one protein to another (90.6%). 22 edges connected natriuretic peptides to plasma proteins, 13 connected echocardiographic variables to proteins, and six linked lipids to proteins. The edges with the highest *d*_*ik*_^*^ score for MACE were the connections between *CAPG* and several proteins including trefoil factor 3 (*TFF3), FSTL3* and ephrin type-A receptor 2 (*EPHA2)*. The highest *d*_*ik*_^*^ scores for the event-free group were between mitogen-activated protein kinase kinase 4 (*MAP2K4*) with prolyl endopeptidase (*FAP*), superoxide dismutase in mitochondria (*SOD2*), histidine-rich glycoprotein (*HRG*) and between plasmin (*PLG*) and vascular endothelial growth factor receptor 2 (*KDR*). Six proteins, *CAPG, FSTL3, EPHA2, TFF3*, tumour necrosis factor receptor superfamily member 19L (*RELT*) and beta-2-microglobulin (*B2M)* were densely connected to many other nodes (i.e. degree above 30). The detailed subnetwork signature is reported in **Supplementary Table 3**.

**Table 3.**
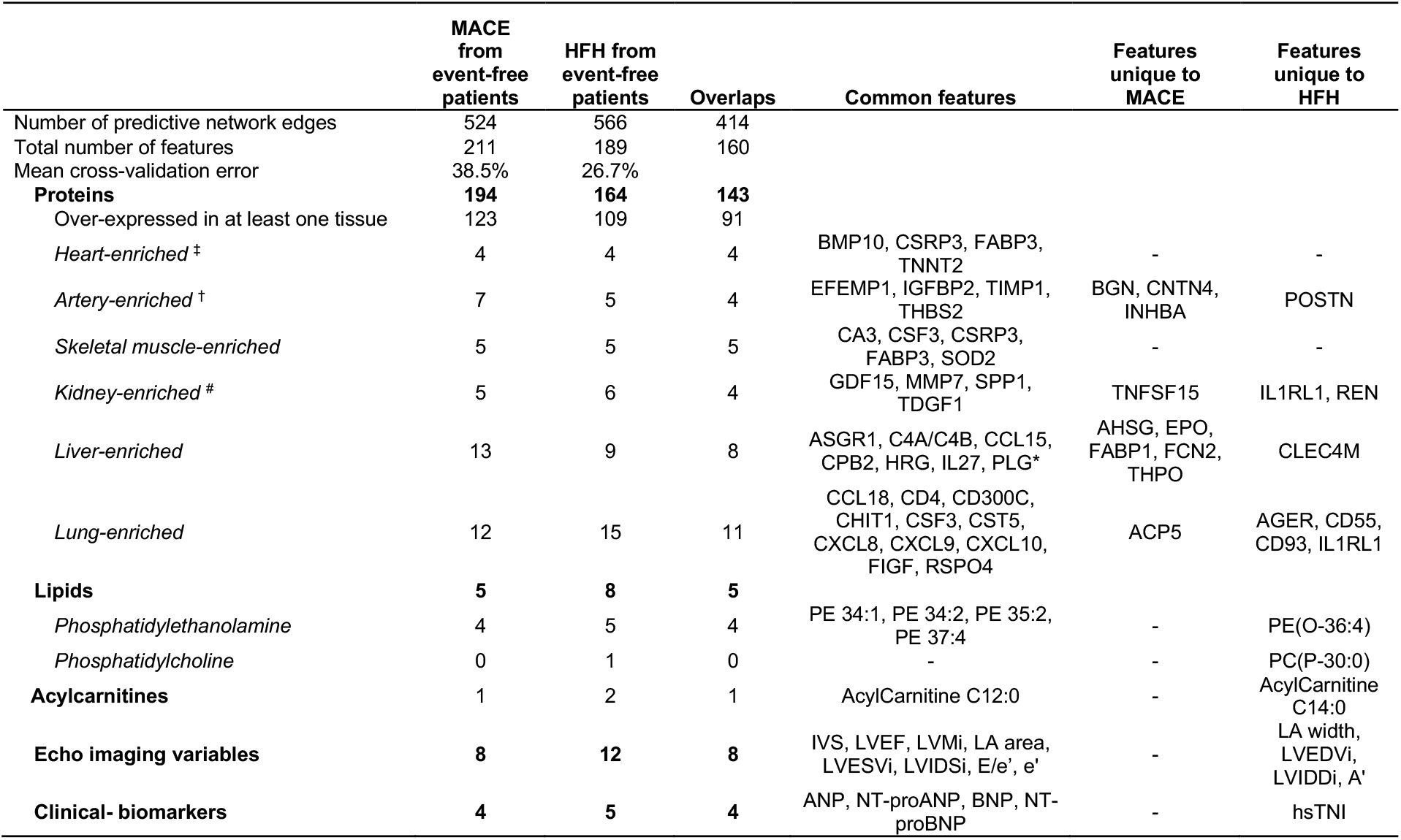
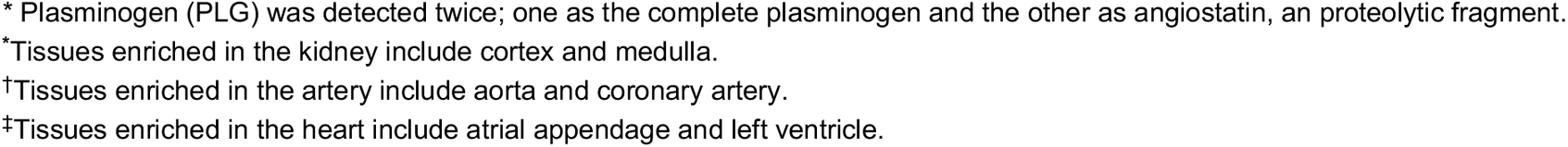
Summary of the predictive subnetwork signatures of lipids, proteins and clinical markers in two comparisons, separating event-free post MI patients from those with a future adverse cardiac outcome (MACE) and from HFH patients.

|  | MACE<br>from<br>event-free<br>patients | HFH from<br>event-free<br>patients | Overlaps | Common features | Features<br>unique to<br>MACE | Features<br>unique to<br>HFH |
| --- | --- | --- | --- | --- | --- | --- |
| Number of predictive network edges | 524 | 566 | 414 |  |  |  |
| Total number of features | 211 | 189 | 160 |  |  |  |
| Mean cross-validation error | 38.5% | 26.7% |  |  |  |  |
| **Proteins** | **194** | **164** | **143** |  |  |  |
| Over-expressed in at least one tissue | 123 | 109 | 91 |  |  |  |
| *Heart-enriched* <sup>‡</sup> | 4 | 4 | 4 | BMP10, CSRP3, FABP3,<br>TNNT2 | - | - |
| *Artery-enriched* <sup>†</sup> | 7 | 5 | 4 | EFEMP1, IGFBP2, TIMP1,<br>THBS2 | BGN, CNTN4,<br>INHBA | POSTN |
| *Skeletal muscle-enriched* | 5 | 5 | 5 | CA3, CSF3, CSRP3,<br>FABP3, SOD2 | - | - |
| *Kidney-enriched* <sup>#</sup> | 5 | 6 | 4 | GDF15, MMP7, SPP1,<br>TDGF1 | TNFSF15 | IL1RL1, REN |
| *Liver-enriched* | 13 | 9 | 8 | ASGR1, C4A/C4B, CCL15,<br>CPB2, HRG, IL27, PLG* | AHSG, EPO,<br>FABP1, FCN2,<br>THPO | CLEC4M |
| *Lung-enriched* | 12 | 15 | 11 | CCL18, CD4, CD300C,<br>CHIT1, CSF3, CST5,<br>CXCL8, CXCL9, CXCL10,<br>FIGF, RSPO4 | ACP5 | AGER, CD55,<br>CD93, IL1RL1 |
| **Lipids** | **5** | **8** | **5** |  |  |  |
| *Phosphatidylethanolamine* | 4 | 5 | 4 | PE 34:1, PE 34:2, PE 35:2,<br>PE 37:4 | - | PE(O-36:4) |
| *Phosphatidylcholine* | 0 | 1 | 0 | - | - | PC(P-30:0) |
| **Acylcarnitines** | **1** | **2** | **1** | AcylCarnitine C12:0 | - | AcylCarnitine<br>C14:0 |
| **Echo imaging variables** | **8** | **12** | **8** | IVS, LVEF, LVMi, LA area,<br>LVESVi, LVIDSi, E/e', e' | - | LA width,<br>LVEDVi,<br>LVIDDi, A' |
| **Clinical- biomarkers** | **4** | **5** | **4** | ANP, NT-proANP, BNP, NT-proBNP | - | hsTNI |

\* Plasminogen (PLG) was detected twice; one as the complete plasminogen and the other as angiostatin, an proteolytic fragment.
\*Tissues enriched in the kidney include cortex and medulla.
†Tissues enriched in the artery include aorta and coronary artery.
‡Tissues enriched in the heart include atrial appendage and left ventricle.

Incorporating endogenous mRNA expression levels of protein coding genes in the heart, arteries, kidneys, liver and lungs (see **Methods**), we carried out biological pathway enrichment of the proteins in our signature for each tissue type, separately. MACE predictive proteins expressed in the heart were related to cell chemotaxis, cell development, extracellular matrix organization, involved in apoptotic signalling pathway, muscle structure development and homeostatic process (**Figure 3B**). Those expressed in the arteries largely overlapped with those expressed in the heart, including cell projection organization, response to hormone, peptidyl-tyrosine phosphorylation and transmembrane receptor protein tyrosine kinase signalling pathway. Proteins specifically expressed in the liver were found to be enriched in inflammatory response and cytokine-mediated signalling pathways, as expected (**Supplementary Table 4**).

**Table 4.** Top ten pairs of data feature with the highest AUC (HFH) and test statistics scores (d_ik_^*^) of the largest magnitude in iOmicsPASS+ in the CDCS and IMMACULATE studies

| Interaction pair | Feature A | Feature B | DataTypeA | DataTypeB | $d_{ik}^*$<br>score<br>for<br>Event-free | $d_{ik}^*$<br>score<br>for HFH | <u>CDCS study</u> | | <u>IMMACULATE study</u> | |
| --- | --- | --- | --- | --- | --- | --- | --- | --- | --- | --- |
|  |  |  |  |  |  |  | AUC | 95% CI for AUC | AUC | 95% CI for AUC |
| EFEMP1 --- NT-proBNP | EFEMP1 | NT-proBNP | Protein | Biomarker | -1.538 | 1.319 | 0.80 | (0.75 - 0.85) | 0.80 | (0.73 - 0.88) |
| CAPG --- FSTL3 | CAPG | FSTL3 | Protein | Protein | -2.206 | 1.891 | 0.79 | (0.74 - 0.84) | 0.69 | (0.59 - 0.79) |
| FSTL3 --- E/e' | FSTL3 | E/e' | Protein | Echo Imaging | -1.690 | 1.449 | 0.78 | (0.73 - 0.83) | 0.73 | (0.61 - 0.85) |
| EFEMP1 --- E/e' | EFEMP1 | E/e' | Protein | Echo Imaging | -2.129 | 1.825 | 0.78 | (0.72 - 0.83) | 0.71 | (0.60 - 0.83) |
| CAPG --- TFF3 | CAPG | TFF3 | Protein | Protein | -2.179 | 1.868 | 0.78 | (0.72 - 0.83) | 0.68 | (0.57 - 0.79) |
| EFEMP1 --- FSTL3 | EFEMP1 | FSTL3 | Protein | Protein | -2.161 | 1.853 | 0.77 | (0.72 - 0.83) | 0.74 | (0.64 - 0.84) |
| RSPO4 --- BNP | RSPO4 | BNP | Protein | Biomarker | -1.469 | 1.259 | 0.77 | (0.72 - 0.83) | 0.77 | (0.68 - 0.87) |
| CAPG --- CST3 | CAPG | CST3 | Protein | Protein | -1.539 | 1.320 | 0.77 | (0.72 - 0.83) | 0.59 | (0.47 - 0.71) |
| EFEMP1 --- GDF15 | EFEMP1 | GDF15 | Protein | Protein | -1.981 | 1.698 | 0.77 | (0.71 - 0.82) | 0.72 | (0.61 - 0.83) |
| CAPG --- B2M | CAPG | B2M | Protein | Protein | -1.963 | 1.683 | 0.77 | (0.71 - 0.82) | 0.61 | (0.49 - 0.74) |

Using the echocardiographic variables and natriuretic peptides as endophenotype, the visualisation of their first-degree neighbours in **Figure 3C** highlights plasma proteins as candidate markers for secondary outcomes with direct implication for cardiac tissue damage and impaired function. All network edges had higher *d*_*ik*_^*^ scores for the MACE group than the event-free group. Among the eight echocardiographic variables in the signature, E/e’ was most connected to plasma proteins, including *EFEMP1, FSTL3, RSPO4*, parathyroid hormone (*PTH*), *THBS2* and *SPON1*, in a decreasing order of *d*_*ik*_^*^ score of MACE. The pair with the highest AUC was the pair between the ratio E/e’ with *EFEMP1*, followed by LV mass with *FSTL3*. BNP was connected with LV mass, LA area, LVEF, LVIDSi and E/e’; ANP was only connected with LV mass. NT-proBNP was connected to key proteins such as *EFEMP1, BMP10, GDF15, IGFB2* and *RSPO4*; NT-proANP was connected to the same proteins except *RSPO4* and to other proteins including *B2M, CAPG, TFF3* and *FSTL3*.

### Subnetwork signature of HFH

Next, we repeated the supervised analysis for HFH, a subset of MACE. iOmicsPASS+ identified 566 edges in the HFH signature (cross-validated misclassification error 26.7%). This subnetwork consists of 164 proteins, 8 lipids, 12 echocardiographic variables and 5 clinical biomarkers, all of which largely overlapped with the MACE signature (**Table 3**). The network with the edges coloured by the signs of partial correlations between connected nodes is visualised in **Supplementary Figure 1B**. Not surprisingly, echocardiographic variables contributed to the signature with more enhanced *d*_*ik*_^*^ scores for the prediction of HFH. In addition to the eight variables in the MACE signature, four additional echocardiographic variables (LA width, LVEDVi, LVIDDi, a’) were included in the HFH signature. hsTNI, seven proteins including *POSTN, IL1RL1, CD55* and *CD93*, two lipids (PE(O-36:4), PC(P-30:0)) and acylcarnitine C14:0 were also unique in this signature.

Similarly, the HFH subnetwork signature showed two contrasting segments, one with higher *d*_*ik*_^*^ scores for HFH (red edges) and the other with lower scores (blue edges) compared to event-free patients (**Supplementary Figure 2A**). Most edges were connections between plasma proteins (84.1%), 34 edges were between natriuretic peptides and proteins, 18 were between echocardiographic variables and proteins, and 12 were between lipids and proteins. Interestingly, the edges with the highest *d*_*ik*_^*^ scores for HFH did not involve NT-proBNP. Instead, *CAPG* connected with *FSTL3, TFF3, B2M* and *EPHA2* and *EFEMP1* connected with *FSTL3, GDF15, B2M, RSPO4, BMP10* and E/e’ (scores between 1.60 to 1.85) had *d*_*ik*_^*^ scores for HFH higher than the score between NT-proBNP with *EFEMP1*. The ratio E/e’, a common measure of diastolic function, was also connected with other proteins such as *FSTL3, RSPO4, THBS1, PTH, SPON1* and the four natriuretic peptides, illustrating its joint prognostic value in predicting future HFH. Full results are reported in **Supplementary Table 5**.

Proteins in the signature with respective mRNA expression levels in the arteries and the heart were enriched for extracellular matrix organization, external encapsulating structure organization and an additional cell chemotaxis in the latter. Kidney-expressed proteins were pro-inflammatory response, lipid response, cytokine production and chemical homeostasis, whereas the proteins of hepatic origin were related to adaptive immune response and cell-growth (**Supplementary Figure 2B**). The results of the tissue-specific enrichment analyses are reported in **Supplementary Table 6**.

Focusing on the the echocardiographic variables, natriuretic peptides and hsTNI, we visualised the direct neighbours of each marker (**Supplementary Figure 2C**). Both ANP and NT-proANP were connected to *BMP10*; both BNP and NT-proBNP were connected to *THBS2, RSPO4, IGFBP2, VEGFD/FIGF* and *SPON1*. Only one protein *IGFBP2* was strongly connected to all four natriuretic peptides. Among the echocardiographic variables, other than E/e’, LV mass connected with all four natriuretic peptides and two proteins (*FSTL3, RELT*). On the other hand, the ratio of LVEF to BNP, LV mass and *TNNT2* (i.e. negative correlation) yielded higher *d*_*ik*_^*^ scores for the HFH group than the event-free group.

### Characterisation of plasma proteins by tissues of origin

To further delineate the proteins directly associated with cardiac assault and tissue damage post-MI, we mapped the proteins to the tissue-enriched genes in the transcriptomic data provided by the Genotype-Tissue Expression (GTEx) database.^18^ Based on our definition of tissue-enriched genes (see **Methods**), 63.4% of the proteins in the MACE signature and 66.5% of the proteins in the HFH signature were enriched in at least one of the 54 tissues catalogued in the GTEx. **Supplementary Figure 3** shows the gene expression of the tissue-enriched markers from the two signatures, illustrating the specificity of hub proteins to the five tissues related to the heart, arteries, kidneys, liver and skeletal muscle.

In the MACE signature, 10.6% of proteins were specifically of hepatic origin, followed by lungs (9.8%) and arteries (5.7%). In the HFH subnetwork signature, the majority were enriched in lungs (13.8%), followed by liver (8.3%) and kidney (5.5%) (**Table 3)**. In both signatures, only four proteins were enriched in the heart tissues: *BMP10, TNNT2*, cysteine and glycine rich protein 3 (*CSRP3*) and hFABP (*FABP3*). Among the proteins enriched in the aortic and coronary arteries, *EFEMP1, IGFBP2, THBS2*, metalloproteinase inhibitor 1 (*TIMP1)*, were shared between both signatures; biglycan (*BGN)*, contactin-4 (*CNTN4)* and inhibin beta A chain (*INHBA)* were unique to the MACE signature, while *POSTN* to the HFH signature. The table of tissue-enriched proteins is reported in **Supplementary Table 7**.

### Validation of subnetwork signatures in IMMACULATE cohort

Lastly, we assessed the predictive power of the two subnetwork signatures in 190 post-MI patients from the IMMACULATE registry, applying the same inclusion criteria. In IMMACULATE, we did not remove any patients with self-reported history of MI due to the small number of MACE cases and the lack of information regarding how long ago the episode took place. This cohort was recruited between 2011 and 2014 in Singapore (median follow-up period 3.9 years). The two cohorts therefore represent post-MI patient populations from two different time periods with substantially heterogeneous ethnic and genetic background and more contemporary clinical management during the follow-up in the latter.

In IMMACULATE, 38 patients had secondary MACE events, of which 23 were HFH, representing a lower frequency of secondary MACE than CDCS, although there may be under-reporting of MACE as hospitalizations due to unstable angina information were not collected. Despite these differences and in the absence of NT-proANP and ANP, the prognostic value of NT-proBNP as a single marker of MACE and HFH remained exceptionally high. **Supplementary Figure 4** clearly shows that NT-proBNP stratifies post-MI patients into three groups of well-separated risks (log-rank test P-values<0.01) based on tertiles in both studies, respectively.

**Figure 4.**
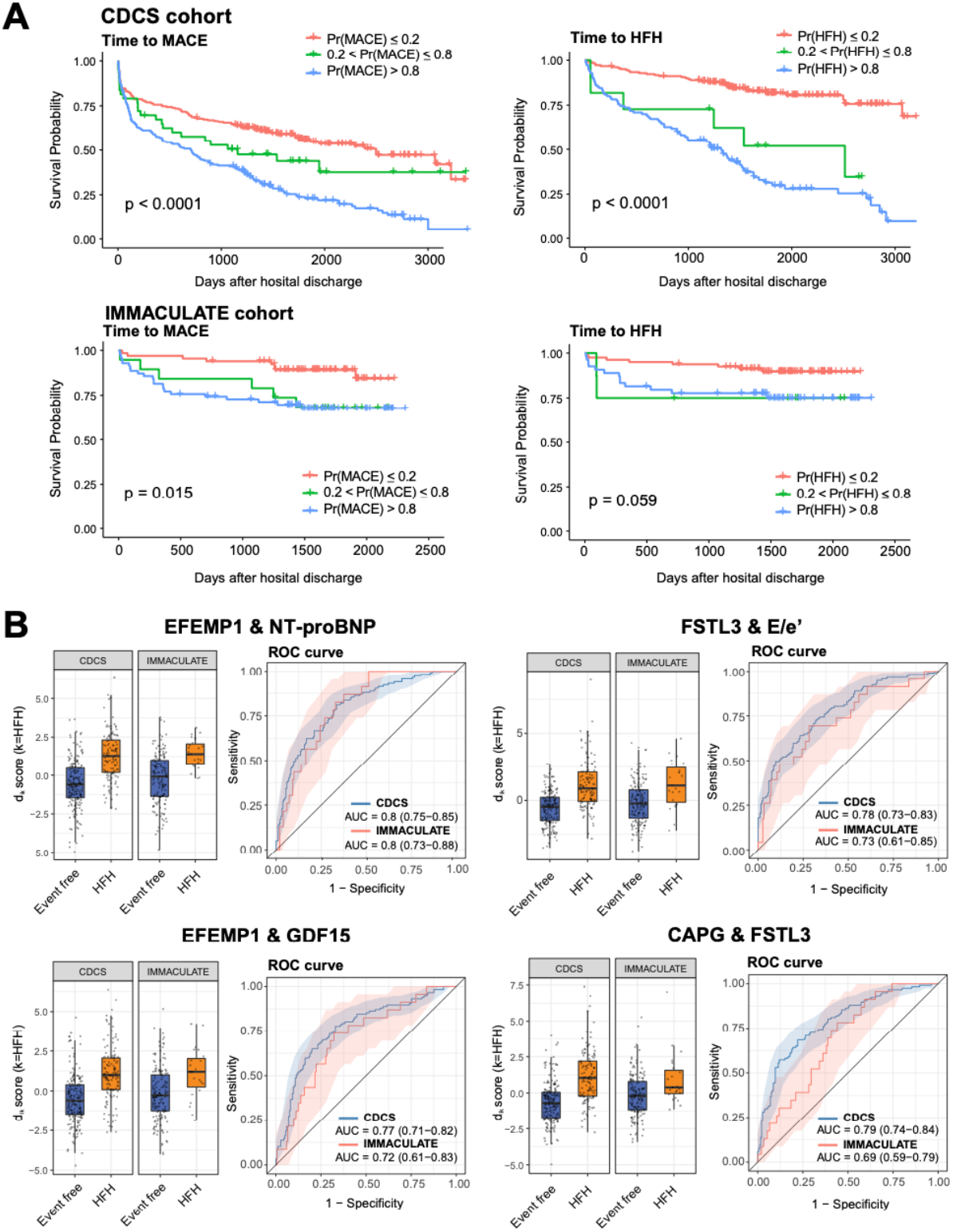
Risk stratification by the patient-level probability scores of the most predictive feature pairs in the CDCS and IMMACULATE cohorts. (**A**) Kaplan-Meier curves with the tertile-based stratification of patients according to the probability scores of MACE and HFH in the training data (CDCS) and the validation data (IMMACULATE). (**B**) Boxplots of the scores of the four top scoring feature pairs predictive of HFH and the respective ROC curves.

Using this signature, we calculated the probability scores of MACE and HFH for the IMMACULATE subjects, a class probability score computed for each patient (see Koh *et al*^*14*^ for details). Stratifying the subjects by the probabilities, we also observed that the survival curves of high and low risk groups were well separated for MACE (P=0.015) and HFH (P=0.059), although the separation of survival curves showed a slightly weaker separation than the results of NT-proBNP as the stratification resulted in more dichotomised grouping of patients (**Figure 4A**). Given that the subnetwork signatures included NT-proBNP, howcver, the lack of improvement in the multivariate signatures alludes to the fact that the global correlation structure of data features is heterogeneous between the two cohorts, and therefore the multivariable signatures of these highly correlated predictors were not directly transferrable between two populations. By contrast, single markers do not have such limitations. For the network signatures to attain improved performance, it would thus require additional tuning of classifier parameters in a specific population with a sufficient amount of training data, i.e. by additional analysis of post-MI patients from Singapore hospitals.

As the patient population differences hindered the portability of predictive subnetworks, we next dissected the class probabilities by smaller units of predictive data features. The linear construction of the logit transform of class probabilities across all edges easily allows us to the contribution of each pair of data features (edge) to the overall class probabilities in a given patient. In particular, we examined the patient-specific, edge-level scores for 41 pairs of data features with absolute group-level *d*_*ik*_* scores above 1.2. Of the 41, **Table 4** shows that the top 10 pairs of data features that had the highest AUC values using their edge-level scores of individual patients. All 11 proteins involved in these connections happened to be network hubs: particularly *CAPG, EFEMP1, FSTL3*, and echo imaging variable E/e’. Similar to the CDCS cohort, we discovered that the differential distribution and the joint predictive powers of most pairs, as measured by AUC of ROC, were comparable to that of NT-proBNP in IMMACULATE. By contrast, other pairs such as the edge connecting *FSTL3* and *CAPG* were not as portable between the two populations (**Figure 4B**). Taken together, a unique subset of subnetwork signatures is validated in an independent cohort and these pairs of data features are as predictive of MACE and HFH as benchmark biomarkers.

## Discussion

In this work, we delineated the global landscape of plasma proteins and lipids in connection with echocardiographic imaging variables and clinical-grade biomarkers in patients hospitalised for a primary acute MI. Plasma proteins carried stronger prognostic signals than lipids in both cohorts, and communities of plasma proteins were associated with increased risks of secondary MACE and HFH. The composite network signatures did not outperform the benchmark biomarkers used in routine care, i.e. natriuretic peptides, in validation in an independent cohort. Notwithstanding the lack of superior performance, our analysis clearly shows that many plasma proteins and echocardiographic variables can stratify the risk of secondary MACE in these patients with prognostic value equivalent to NT-proBNP, and the findings may also point to pathophysiological mechanisms reported by plasma proteins.

Hub proteins in both predictive subnetworks were *CAPG, EFEMP1*, and *FSTL3. CAPG*, referred to as Cap-G, is a Ca^2+^-sensitive actin binding protein in the gelsolin/villin family of barbed end blocking proteins^19^, notably expressed in macrophages. Given the ambivalence between the pro-inflammatory role of bone marrow / spleen-derived, classically activated macrophages during the early inflammatory phase of the primary MI and the cardioprotective role of resident cardiac macrophages during the repair stage^20^, the elevation of *CAPG* levels in these patients indicates a shift in the macrophage population following the primary thrombotic occlusion. *EFEMP1*, also known as fibulin-3, is an extracellular matrix glycoprotein implicated in vascular endothelium remodeling,^21^ and plays a role in reducing vascular calcification and inhibiting metalloproteinases in oxidative stress.^22-24^ While fibulin-3 does not seem to be a direct mechanistic cue in cardiac repair, we observed strong predictive signals of secondary events with moderate correlations with NT-proBNP. *FSTL3*, an extracellular regulator of TGF-β family cytokines such as activin A, is involved in various biological functions including cell proliferation and inflammation, and altered transcriptomic regulation of the gene with another follistatin family member *FSTL1* in myocardium has been associated with HF severity.^25^ This protein also showed positive partial correlation with diastolic function variable (E/e’). *FSTL3* was also characterised as a stress-induced regulator of cardiac hypertrophy through Smad signalling pathway modulation in a mouse model^26^, opening up an opportunity for investigation as a therapeutic target for HF.

In this study, circulating lipids showed weak prognostic values for MACE in both cohorts. While ceramides have been reported to be useful in the assessment of CVD risk,^27-29^ none of the ceramides were significantly different between MACE and event-free patients. When we investigated the lipid-based CERT1, CERT2, SIC risk scores, and the recently proposed cardiac lipid panel consisting of triacylglycerol (TG 18:1/18:0/18:0), PC 16:0/18:2 and sum of three isobaric SMs (d18:1/23, d18:2/23:0, d17:1/24:1) for predicting chronic HF,^30^ none of those lipids in our panel were significantly different across the different outcome (data not shown).

Although lipids did not show a comparable prognostic power to proteins, both subnetwork signatures included four lipid transport proteins, namely *APOL1, APOE, APOA1* and *FABP3. FABP3* encodes hFABP, a marker of myocardial injury, and it was connected with multiple acylcarnitine species in both subnetwork signatures. hFABP has previously been implicated in HF, where reduced fatty acid utilization in the heart leads to the progression of chronic heart failure, left ventricular hypertrophy and remodelling.^31, 32^ Interestingly, here we found that circulating levels of hFABP and acylcarnitines (C12:0, C14:0, C14:1, C14:2, C16:1) were jointly increased in MACE and HFH patients, suggesting that the levels in circulation may reflect leakage products from the remaining cardiac injury.

Taken together, our multi-modal data integration approach underscores the importance of effective integration of multi-omic measurements, cardiac imaging variables, and clinical biomarkers. Although univariate assessment of individual marker candidates is of utmost importance for biomarker discovery, a plethora of molecular features representing biologically interpretable signals may go unnoticed as false negatives when the underlying correlation structure is not carefully explored. As blood is frequently the only available reporter tissue in CVD biomarker discovery, we also annotated individual proteins in terms of the potential tissue(s) of origin. With the emergence of similar large-scale, blood plasma-based investigation of other diseases, we expect this and other novel data analytic approaches to play important roles in the prioritization of potential therapeutic targets.

## Data Availability

iOmicsPASS+ is publicly available as an open-source R package in the GitHub repository (https://github.com/cssblab/iOmicsPASSplus). The proteomic and lipidomic data in support of the findings of this study is available at https://github.com/Hiromikwl/Data_iOP. Patient-level clinical records, including echocardiographic imaging variables and clinical biomarkers, can be shared after reasonable preliminary discussion and agreement with both corresponding authors due to ethics and privacy concerns. 

## Acknowledgment

This work was supported in part by grants from National Medical Research Council of Singapore (CIRG17may014 and MOH-000280 to MYC), from New Zealand: The Coronary Disease Cohort Study was funded by the Health Research Council of New Zealand (Program Grants 02/152, 08/070, 11/1070); National Heart Foundation of New Zealand; New Zealand Lotteries Grant Board; Foundation for Research, Science and Technology, and the Christchurch Heart Institute Trust, and National Medical Research Council of Singapore (to AMR). AMR holds the NZ Heart Foundation Chair of Cardiovascular Studies; the National University of Singapore via the Life Sciences Institute (LSI) (to MRW), National Research Foundation (NRFI2015-05, NRFSBP-P4 to MRW) and A*STAR (I1901E0040 to MRW); Singapore Ministry of Education (MOE2016/T2/1/001, NMRC/CG/M009/2017, MOE2018/T2/2/058, MOE-T2EP20121-0018 to HC).

## Methods

### The Coronary Disease Cohort Study (CDCS)

The CDCS cohort consists of 2,140 patients recruited from two tertiary hospitals (Christchurch Hospital and Auckland City Hospital) in New Zealand (NZ) for an acute coronary syndrome (ACS) event from 2002 to 2009. CDCS was a registered trial - ACTRN 12605000431628. Participants were invited to return to the hospital 30 days after discharge for baseline measurements. Patients were excluded from the study if their life expectancy was estimated as less than 3 years. The study was approved by the New Zealand Multi-region Ethics Committee (CTY/02/02/018) and all participants gave written informed consent. More information on the study can be found in Prickett *et al*.^33^ Details of the cohort, and the subset of 741 patients analysed in this report can also be found in **Supplementary Information**, along with further inclusion and exclusion criteria for the supervised analysis (N=464).

### The Improving Outcomes in Myocardial Infarction through Reversal of Cardiac Remodelling (IMMACULATE) cohort

The IMMACULATE cohort consists of 859 patients who were admitted for MI into three local hospitals in Singapore (National University Hospital, Tan Tock Seng Hospital and Singapore General Hospital) from 2011 to 2014. The patients were followed up for a median of 2.0 years (interquartile range IQR: 1.9-2.1 years) from their hospital discharge date. The study was approved by the institutional review board and the ethics committee at Singapore’s National Healthcare Group Domain Specific Review Board (DSRB 2013/00248 and 2013/00635). All participants provided written informed consent. Criteria for initial inclusion in IMMACULATE and the selection of 190 patients analysed in this report can be found in **Supplementary Information**.

### Proteomics, lipidomics and clinical biomarker measurements

Protein abundance was measured using Slow Off-rate Modified Aptamer (SOMAmer)–based capture array, called SOMAscan (somaLogic, Inc, Boulder, CO, USA).^34^ Targeted lipidomics experiments were performed using an Agilent 6495A triple-quadrupole (QQQ) mass spectrometer coupled to an Agilent 1290 Infinity-II UHPLC system, with automated data processing and quality control by the MRMkit tool.^35^ Acylcarnitines were also measured as part of this panel. Well-established cardiac markers, including natriuretic peptides (ANP, BNP, NT-proANP, NT-proBNP), high sensitivity troponin-I (hsTNI) and creatinine, were measured using clinical-grade assays. Due to the higher accuracy and sensitivity of the biomarker assays, we removed four corresponding proteins measured in SOMAscan that were targeting the same proteins to prevent duplicated information. Refer to **Supplementary Information** for full details on the assay platforms and data processing.

### Echocardiography

Standard M-mode measurements of LV dimensions, wall thickness, LA dimensions were made according to the recommendations of the American Society of Echocardiography (ASE).^36^ LV volumes and the derived left ventricular fraction was measured by the Simpson modified biplane method.^37^ Doppler indices were also acquired according to the ASE guidelines for Doppler echocardiography.^38^ Mitral pulsed-wave Doppler velocities of early passive (E) and atrial (A) filling were obtained from the apical 4-chamber view with a 5 mm sample volume placed between the tips of the mitral leaflets,^39^ and systolic (S), diastolic (D), and atrial reversal (A) pulmonary vein velocities were acquired from the apical 4-chamber view with a 5 mm sample volume placed 1 cm into the right upper pulmonary vein. Lastly, Ommen *et al*. provides the methods of tissue Doppler measurements of early diastolic velocity (e’), late diastolic velocity (a’), and systolic velocity of the myocardial muscle (s’), where we used the average of the septal annulus velocity and the lateral annulus velocity for each measurement.^40^ Echocardiographic variables with missing data proportion less than 25% were considered. This filter resulted in 19 echocardiographic variables to be considered for the downstream analysis. Missing values in those variables were imputed together with missing entries with the six clinical biomarkers using multiple imputation by chained equations (MICE).^41^

### Ascertainment of MACE

The primary outcome of interest was 5-point MACE, defined as a composite outcome that includes non-fatal acute MI, non-fatal stroke, hospitalization for unstable angina, heart failure and/or a cardiovascular (CV)-related death.^42, 43^ Stroke included the occurrence of an ischemic stroke, haemorrhagic stroke and transient ischemic attack (TIA). Acute MI included both ST-elevation (STEMI) and non ST-elevation (NSTEMI) myocardial infarction. Diagnosis of MI and HF adhered to established clinical guidelines on the diagnosis and management of MI and HF.^44-49^ Adverse remodelling of the heart was assessed by changes in echocardiographic left ventricle ejection systolic volume (LVESV) from the baseline visit to first follow-up at four months.

All patients were followed from the time of their primary hospital discharge to a future major adverse cardiac event, death or end of study, whichever was earlier. For both CDCS and IMMACULATE, we defined three phenotypic outcomes: (1) patients who remained event-free (event-free), (2) patients with 5-point MACE (or MACE) and (3) patients hospitalised due to HFH. Patients with HFH are a subset of those with MACE and are considered as HFH regardless of the precedence of any MACE outcome. Event-free individuals are those who remained free of any major cardiac event (i.e. acute MI, stroke, unstable angina, HF, CV-related death) during the entire course of the follow-up period. To prevent confounding in the predictive analyses, we removed patients with previously remodelled hearts and those with a medical history of stroke or HF; and among those with a history of prior MI we retained only those in whom this event had occurred more than five years prior to recruitment. We use the term MACE to refer to the traditional 5-point MACE throughout this paper unless otherwise stated and 4-point MACE to refer to the subset of MACE that excludes HFH patients.

### Statistical analysis

Clinical characteristics of the CDCS cohort were compared across event-free patients and patients with MACE. For continuous variables, two-sample *t*-test was used to compare the difference in means across groups, whereas for categorical variables, Chi-squared test was used to test for associations with groups. Fisher-exact test was used when any of the cell-frequencies were less than five. For variables with missing or unknown self-reported entries, they were removed before performing the statistical test for association. All P-values were adjusted for multiple testing correction using the Benjamini-Hochberg’s (BH) method to control the false discovery rates (FDR) below 0.05. The AUC of the ROC was also computed for each marker to evaluate their predictive performance.

For the estimation of a confounding-free partial correlation-based network, we utilised all the non-missing markers across 741 CDCS samples and computed a cross-covariance matrix using all pairwise complete observations as input to graphical LASSO^13^, for the inference of the network structure and calculation of partial correlations, built in as a module within iOmicsPASS+. Note that the partial correlations reported in this work are regularised values, not unbiased estimates. Using the estimated network, we searched for sparse predictive subnetwork signatures across 464 patients to differentiate (1) MACE patients from event-free one, as well as separate (2) patients with HFH from event-free ones, with the assistance of 10-fold cross-validation (CV) for parameter optimisation. The network signatures are characterised by group-specific centroids of edge-level scores. For every *i*-th edge (pair of correlated data features), interaction scores were calculated for individual patients and modified test statistics (scaled group centroids), denoted by *d*_*ik*_*, were computed for all groups (*k* = 1, …, *k*). The latter represents whether the two connected nodes had higher or lower values in the same or opposite directions in the *k*-th phenotypic group, depending on the sign of the respective partial correlations. These test statistics were penalised iteratively to select the optimal sparse networks for the two classification tasks, and were reflected in the colour and thickness of edges in the network visualisations. Details of this method are described in Koh *et al*.^14^

The proteins included in the final predictive signatures were annotated in terms of their biological processes in Gene Ontology (GO) using gProfiler^50^. Enrichment analysis was repeated with an additional filtering of query and background gene lists with regard to the minimal expression level (transcript per million reads, or TPM > 5) in each tissue type as provided by the GTEx.^18^ The tissue types considered in this study include the heart (atrial appendage, left ventricle), arteries (aorta, coronary), kidneys (cortex, medulla), liver, lungs and skeletal muscle.

All the aforementioned analyses were carried out in R studio (R version 4.0.2),^51^ using external R packages huge,^52^ gplots,^53^ mice,^54^ pROC,^55^ clusterProfiler,^56^ as well as iOmicsPASS+ presented in this paper. Visualization of networks were done using Cytoscape (version 3.8.2).^17^ The full description of statistical analysis and network-oriented classification analysis using iOmicsPASS+ is provided in **Supplementary Information**.

