## Supplementary Information for "Plasma multi-omic and cardiac imaging network signatures predict poor long-term outcomes after acute myocardial infarction"

### Supplementary Figures

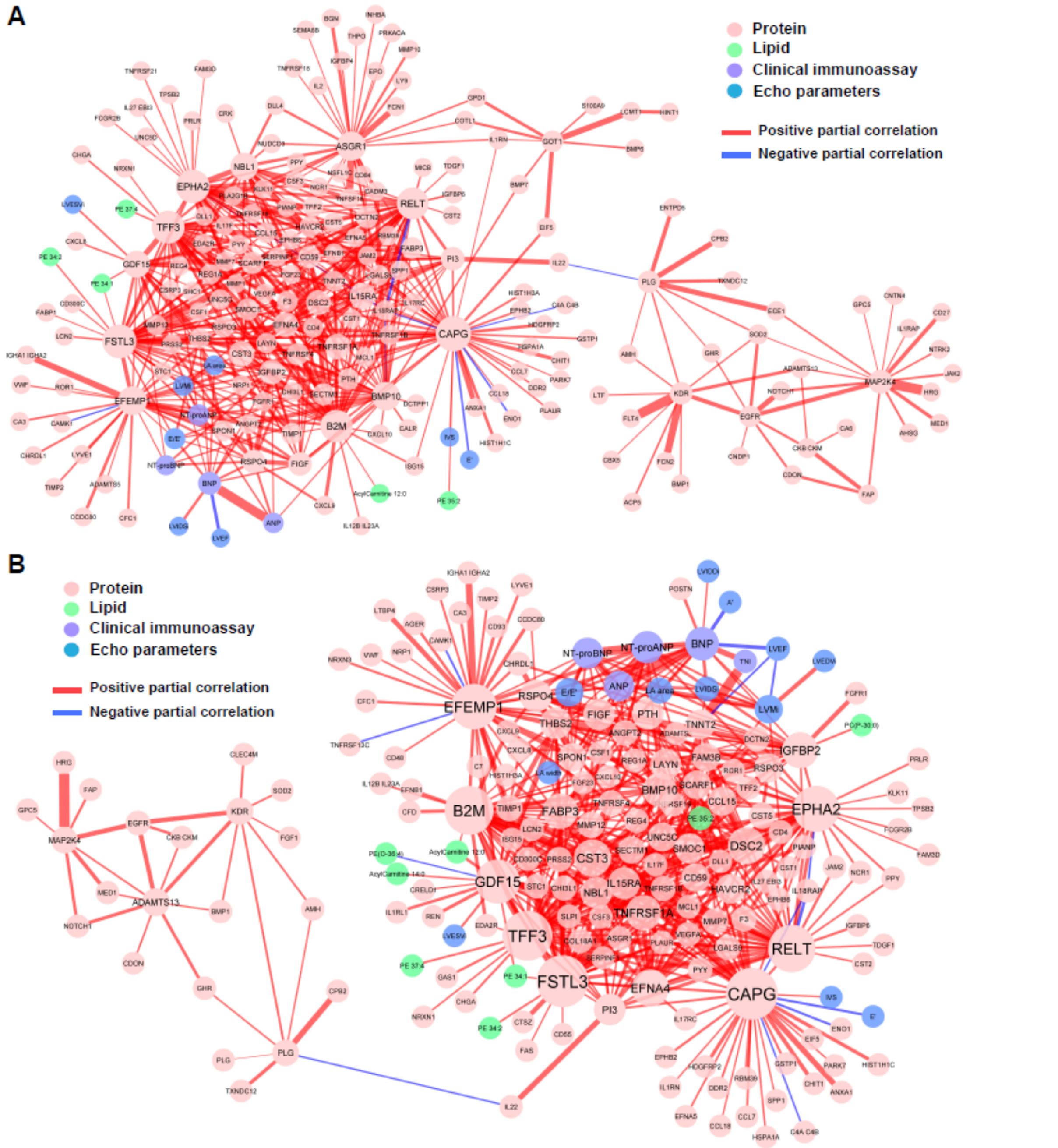


**Supplementary Figure 1.** Network of the plasma proteins, lipids and acylcarnitines, echocardiographic imaging variables, and clinical biomarkers in the (**A**) MACE signature and the (**B**) HFH signature that mirrors the same network in **Figure 3A** and **Supplementary Figure 2A**. This time, the edges were coloured according to the sign of partial correlations (red for positive and blue for negative), where most connected nodes were positively correlated. The thickness of the edges reflects the absolute magnitude of the partial correlation.

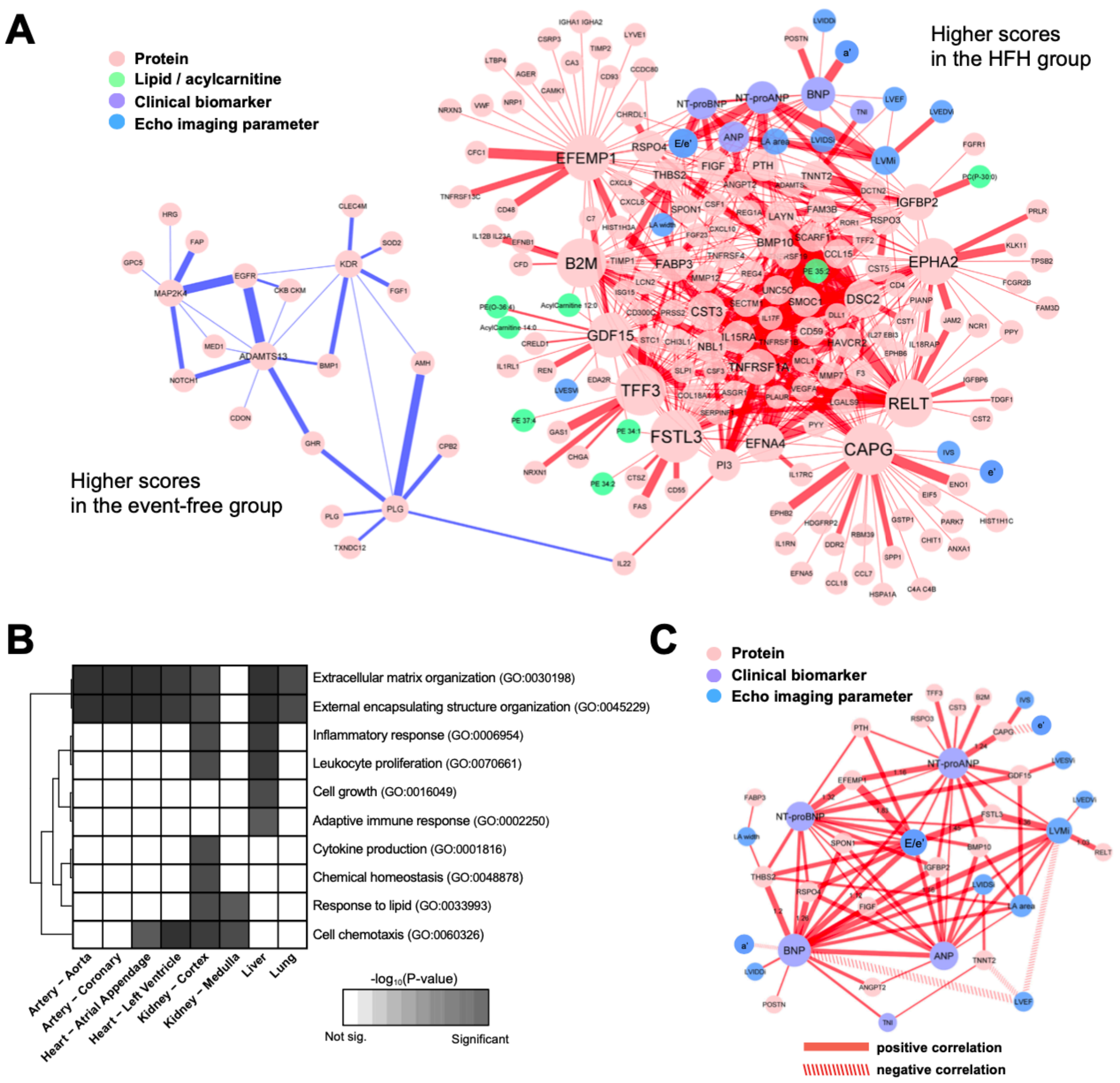


**Supplementary Figure 2.** (**A**) The HFH subnetwork signature. The edges were coloured by the sign of the edge-specific scores for the HFH group (positive in red, and negative in blue, respectively). The thickness of edges reflects the magnitude of the scores. (**B**) Enrichment of biological functions in the constituent nodes in the panel (**A**) accounting for tissue-specific expression of genes at the mRNA level. (**C**) A subset of the HFH signatures consisting of first-degree neighbours of echocardiographic variables and clinical biomarkers, with line properties of the edges set according to the signs of respective partial correlations (positive in solid, and negative in dashed).

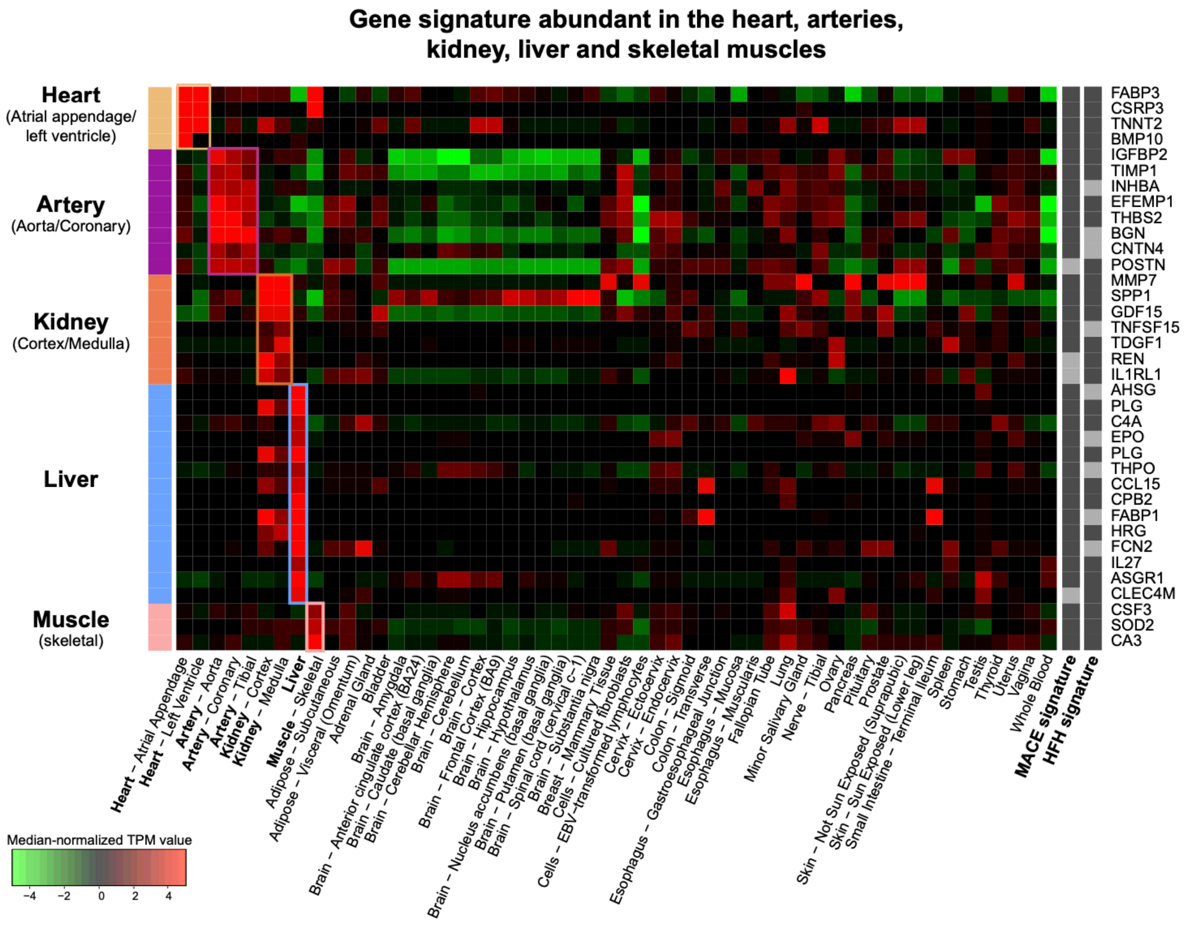


**Supplementary Figure 3.** Tissue specific gene expression (median-normalised TPM values) of 36 plasma proteins in the MACE and HFH signatures with specific enrichment in heart, arteries, kidneys, liver, and skeletal muscle according to the GTEx database.

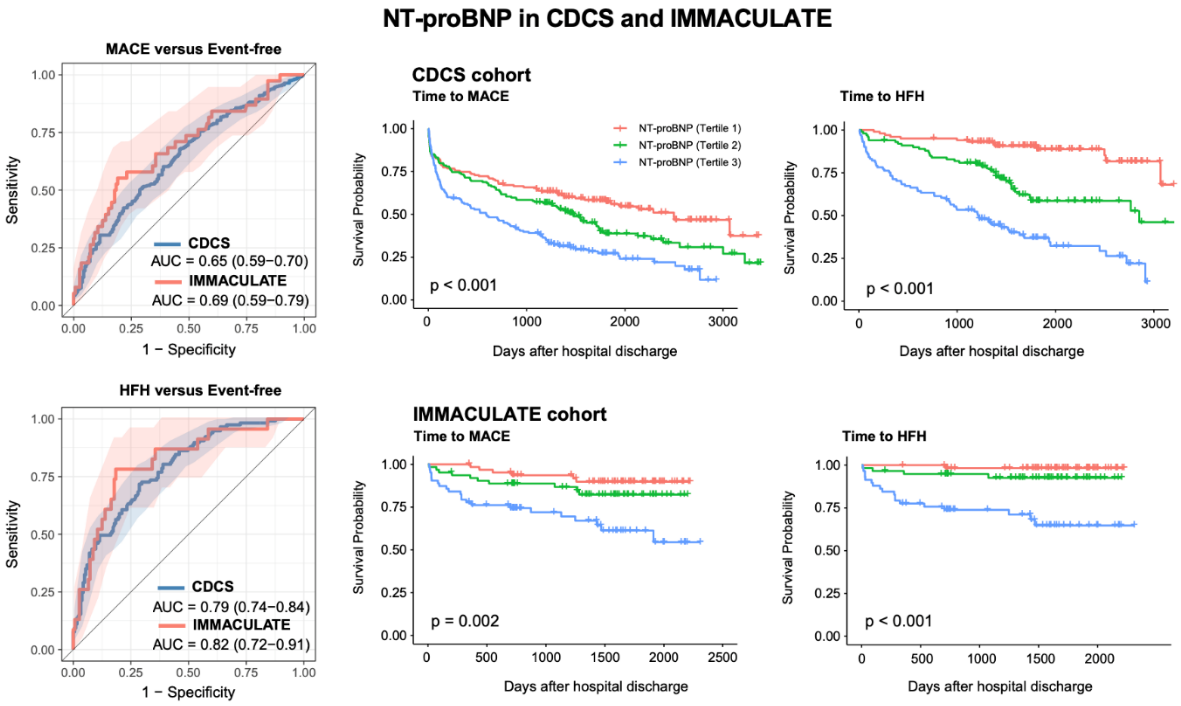


**Supplementary Figure 4.** The prognostic value of NT-proBNP for MACE and HFH in the two cohorts, measured by the ROC analysis as well as the separation of risk profiles via tertile-based stratification.

### Supplementary Tables

**Supplementary Table 1.** Hypothesis testing (two-sample t-test) of data features between MACE and event-free patients and between the HFH subgroup and event-free patients in the CDCS cohort.

**Supplementary Table 2.** Partial correlations associated with the edges connecting at least one of the echocardiographic variables and clinical biomarkers in the CDCS cohort.

**Supplementary Table 3.** MACE signature derived by the supervised analysis module of iOmicsPASS+ using the CDCS data, with edge specific scores of the two groups (MACE and event-free).

**Supplementary Table 4.** Biological function enrichment analysis of the proteins in the MACE signature, accounting for tissue-specific mRNA expression.

**Supplementary Table 5.** HFH signature derived by the supervised analysis module of iOmicsPASS+ using the CDCS data, with edge specific scores of the two groups (HFH and event-free).

**Supplementary Table 6.** Biological function enrichment analysis of the proteins in the HFH signature, accounting for tissue-specific mRNA expression.

**Supplementary Table 7.** Summary table of tissue-specific over-expression of individual genes encoding the plasma proteins at their respective, potential sites of origin.

#### The Coronary Disease Cohort Study (CDCS)

The CDCS cohort consists of 2,140 patients admitted into two tertiary hospitals (Christchurch Hospital and Auckland City Hospital) in New Zealand (NZ) for an ACS event, recruited from 2002 and 2009. They were invited to return to the hospital one month after discharge for baseline measurements. Patients were excluded from the study if they had a severe comorbidity that limited their life expectancy to less than 3 years. Clinical measurements, blood sample collection, and echocardiogram were all obtained at baseline (30 days from discharge) and 4 months, 6 months and 12 months after discharge. These individuals were followed up until the end of the study on 31st January 2012, death or lost-to-follow-up. Subsequent clinical events and mortality information were obtained from the NZ Nation Health Information System. The study was approved by the New Zealand Multi-region Ethics Committee (CTY/02/02/018) and all participants had provided a written informed consent prior to their participation. More information on the study can be found in Prickett *et al*[1].

Out of these 2140 patients, a nested sub-cohort of 750 patients matched by age and gender was selected, and both aptamer-based SOMALOGIC protein assay and multiple reaction monitoring (MRM)-based lipidomics were used to measure proteins and lipids, respectively. Among these, 250 patients were readmitted for HF after their hospital discharge, 250 patients were readmitted for myocardial infarction (MI) episode and 250 patients were selected as controls without any recurrence of MI or incidence of heart failure hospitalisation (HFH). The controls were further defined as free of adverse cardiac remodelling and any cardiovascular-related death over the follow-up period. They were selected by matching each HF patient to a MI patient and a control by age and gender using the MatchIT package in R[2].

Following a quality control step for the molecular data, 741 patients were retained for network inference. For the supervised analysis using the iOmicsPASS method, 286 patients were removed due to the following exclusion criteria: 69 patients with cardiac remodelling, 81 patients with a medical history of heart failure, 97 patients with history of stroke, 78 patients with a prior MI within the previous five years, and 19 patients with missing or unknown status to minimise possible error in the outcomes. A total of 464 patients was retained for predictive subnetwork identification for MACE and HFH. Of those,185 patients remained event-free, and 279 patients had a secondary MACE, including 117 patients hospitalised for HF during follow-up.

#### The Improving Outcomes in Myocardial Infarction through Reversal of Cardiac Remodelling (IMMACULATE) registry

The IMMACULATE registry consists of 859 patients who were admitted for MI into three local hospitals in Singapore (National University Hospital, Tan Tock Seng Hospital and Singapore General Hospital) from 2011 to 2014. The patients were followed up for a median of 3.9 years (interquartile range IQR: 2.0-4.8 years). Inclusion criteria were patients with a ST-segment elevation MI (STEMI) or non ST-segment elevation MI (NSTEMI) with ischemic chest pain or angina symptoms, concentrations of troponin exceeding $99^{th}$ percentile URL and >50% occlusion of one or more coronary arteries based on angiography. Patients were excluded if they were anaemic (haemoglobin<8g/dl in men and <7g/dL in women), had severe renal impairment (estimated glomerular filtration rate, eGFR<15 ml/min/$m^2$), were in cardiogenic shock not able to be weaned off inotropes or intra-aortic balloon pump, history of malignancy or other conditions limiting life expectancy diagnosed within the last 12 months.

Hospital records were used to capture readmissions due to a recurrent ACS event or other morbidity. Mortality data and primary cause of death was obtained from the National Registry of Death Office (NRDO). The study was approved by the institutional review board and the ethics committee at Singapore's National Healthcare Group Domain Specific Review Board (DSRB 2013/00248 and 2013/00635). All participants provided written informed consent.

We selected 200 IMMACULATE patients with clinical information and blood samples obtained at three time points: (1) within 24-72 hours of admission, (2) 1 month after hospital discharge and (3) 6 months after hospital discharge. At all three time points, plasma lipids and proteins were profiled and echocardiography undertaken. In the current report, the IMMACULATE study serves as validation and only measurements one month from hospital discharge post-MI, a temporal match with CDCS, have been used. Among these, 190 subjects had all relevant biochemistry data for cardiac markers, proteomics, and lipidomics data. We further removed one patient with self-reported history of HF and six patients with history of stroke. However, we did not remove any patients with self-reported prior MI as the duration of prior MI to primary ACS event was unknown, as well as due to the constraint in the sample size. As a result, we recorded 152 patients who remained event-free and 38 patients with secondary 5-point MACE (15 4-point MACE and 23 HFH). Here, we use IMMACULATE cohort as a validation for the evaluation of classification performance.

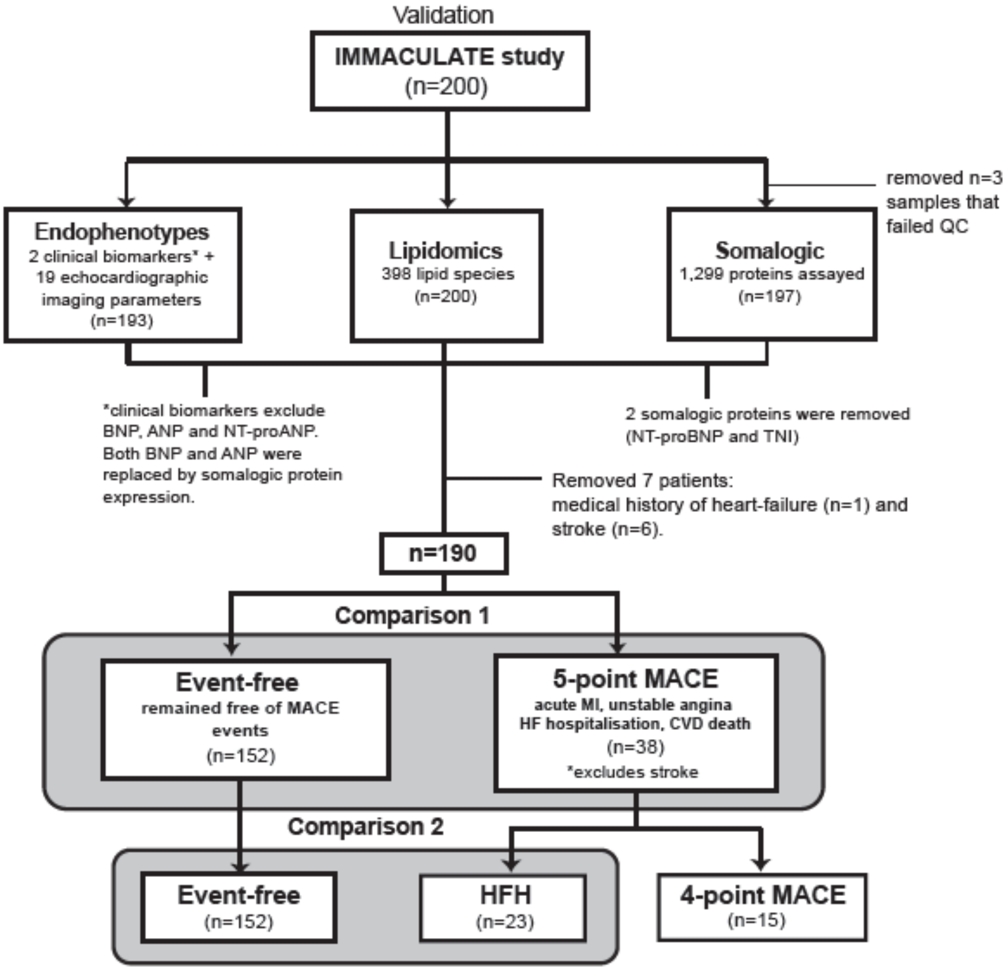


**SOMALOGIC Protein Assay**

Plasma samples (50 µL) collected 30 days after the index event were analysed using a Slow Off-rate Modified Aptamer (SOMAmer)–based capture array, "SOMAscan" (somaLogic, Inc, Boulder, CO, USA)[3]. A total of 1,305 human proteins were quantified (47% secreted proteins, 28% extracellular domains and 25% intracellular proteins). Details of the experimental methods and data analysis protocol are described in Chan *et al*.[4] We note that the protein names in all diagrams and tables are given by the gene symbols of the encoding genes (all italicised), with the exception of NT-proBNP and cardiac troponin I, which are measured by both immunoassay and SOMALOGIC.

**Lipidomics Assay**

Samples were randomised into 10 analytical batches and batch quality control (BQC) samples were prepared by pooling equal plasma aliquots from all the samples. To assess analytical performances, one BQC sample was included for every ten study samples. Plasma (10 µL) was mixed with 190 µL 1-butanol/methanol (1:1, v/v), [BuMe], containing 4.5 µL of SPLASH™ Lipidomix® Mass Spec Standard I (#330707) and 4.5 µL Cer/Sph Mixture I (#LM6002) from Avanti Polar Lipids, Inc. The mixture was vortexed for 30 seconds, then sonicated for 30 min (ice was added into the sonication bath) and then centrifuged at 14,000 x g for 10 min at 10°C. The supernatant was transferred into autosampler vials for mass spectrometry. Extracted blanks were prepared using the same extraction protocol but without plasma samples. Technical quality control (TQC) samples were generated by pooling lipid extracts from the study samples and were used to monitor the analytical system performance.

Analysis of plasma extracts was performed on an Agilent 6495A QQQ mass spectrometer coupled to an Agilent 1290 Infinity-II UHPLC system and the MRM data acquired in positive ion mode were processed using the MRMkit tool.[5] Peaks were integrated and the concentration data were further normalised by a time-trend and batch effect correction algorithm built in MRMkit. Lipid species with the overall coefficient-of-variation smaller than 25% were included for the downstream analysis.

**Quality control for proteomics and lipidomics data**

For both CDCS and IMMACULATE studies, only the measurements collected one month after hospital discharge (baseline) were used. For the CDCS study, a total of 376 unique lipids were used for the subsequent analysis. For the IMMACULATE study, a total of 418 lipid species were retained for validation analysis for relaxed criteria (CoV 50%). Internal standards were used to normalise the raw peak areas in the corresponding lipid class and concentrations were further normalised to the protein concentration in the original sample. Due to the methodology used in our study, we do not report absolute concentration values. Endogenous species were quantified using one standard per lipid class, hence our method can only deliver relative quantification results, which does not hamper testing of concentration differences between subjects within any given lipid species. For the proteome analysis in both studies, a standard SOMALOGIC 1.3k array was used. For the purpose of the analysis, we removed 5 proteins that were from human viruses (i.e. human papillomavirus) and samples that failed QC, resulting in 747 samples in CDCS and 197 in IMMACULATE study.

### iOmicsPASS+

iOmicsPASS has been expanded and reimplemented as iOmicsPASS+ to allow for greater flexibility and generalizability to other types of omics data. First, the tool now allows for only one network and one data as input. If only one data (X) is used as input, then the network input is assumed to be one which links the features within the data. If more than one data is used as input (e.g. X, Y and Z), then a second network should be provided to link the features between the data in X to the data in Y. Data Z is used to normalise against matching features in data Y as in the original iOmicsPASS.

Second, the network input now allows for direction of association for the pair of interacting or co-varying data features. The user can specify an additional column in the network file to include whether the association between the pair of features is positive (i.e. 1) or negative (i.e. -1). For the latter, the interaction scores are calculated as ratios (differences of two Z-scores), instead of the usual product (sum of two Z-scores). Within iOmicsPASSv2plus, a sample correlation matrix is first calculated to determine if the direction of the association is consistent with the indicated direction by the user in the network. An edge with interaction score is created only if the sign of the correlation between two data features is consistent with the specified direction of association. Otherwise, if inconsistent, the pair of data features is not used as predictive features for the subsequent analysis. This enables the application to other omics data such as microRNAs and DNA methylation where the inhibition of gene expression occurs as a result of interaction with the target genes.

Last, several new modules are added to the R package to facilitate users to build, install, create input parameter file and run the software. One of the most important module is a network inference module which estimates the best co-varying network between the different types of data, using an existing R-package "Huge"[6], which carries out graphical LASSO (GLASSO) estimation. Within the module, we propose two different approaches for estimating the network: a supervised approach and a hybrid approach. In the supervised approach, the network is fully estimated from the precision matrix while the latter fuses a user-provided network input based on prior knowledge with the estimated GLASSO-derived partial correlation network to produce a combined network.

Another essential module in the R package is the prediction module that uses the iOmicsPASS results to assign new samples to the phenotypic groups based on the set of predictive signatures. The prediction module uses a discriminant model to assign each sample to the phenotype group with the highest discriminant score. In this module, the discriminant score is modified such that the prior probability incorporates other clinical information such as age, gender and BMI, that may influence the classification.

The following sections will describe the network inference method in greater detail.

**Notation**

We first define a few notations for the subsequent sections. Consider a *p*-dimensional data, $X = \left(X_1, X_2, \dots, X_p\right)^T$, with mean vector μ and covariance matrix $\Sigma_{p\times p} = \left(\sigma_{ij}\right)_{1\le i,j\le p}$.

Let

$$X_i = (X_{1i}, X_{2i}, \dots, X_{ni}),\ i = 1, \dots, p$$

denote cross-sample observations of data feature $i$, where $n$ is the sample size of the data. The covariance matrix $\Sigma$ is estimated by the sample covariance matrix $S$, where the $(i,j)^{th}$ entry represents the sample covariance of $X_i$ and $X_j$ calculated as:

$$s_{ij} = \frac{1}{n-1}\sum_{r=1}^{n}(x_{ri} - \overline{x}_i)\left(x_{rj} - \overline{x}_j\right) \quad (1)$$

Then, the precision matrix, $\Omega = (\omega_{ij})_{1 \le i,j \le p}$, is the inverse of the covariance matrix $\Sigma$ and the $(i,j)^{th}$ entry describes the conditional dependency of $X_i$ and $X_j$ on all other (*p*-2) variables If $\omega_{ij} = 0$, then $X_i$ and $X_j$ are said to be conditionally independent given other variables present. If $\omega_{ij} \neq 0$, then $X_i$ and $X_j$ are not conditionally independent and an edge can be form between the two in a network setting.

### Network inference module

In this module, there are two proposed ways to create a pseudo network, linking one –omics to another. First, a supervised approach is completely driven by the estimates from the data. Second, a hybrid approach, which uses a mix of what is known as a prior and what is derived from the data by the supervised approach to produce a combined network adjacency matrix. The subsections below provide a detailed description of the method in the two approaches.

#### *Supervised approach*

In the supervised approach, the input data is first standardised to unit standard deviation so that the sample covariance matrix and the sample correlation matrix are equivalent. Before the sample covariance matrix is computed, PCA is used to identify any outliers, defined to be more than 4 SDs away from the median of the first 2 PCs derived from concatenating all the data into a single matrix. After filtering outliers, the sample covariance matrix $S$ is computed and corrected to the nearest PSD matrix if any of its eigenvalues are negative.

To convert a matrix $A$ to the nearest PSD matrix, the approach by Nicholas J. Higham [7] is used:

$$A_{PSD} = \frac{B+H}{2} \tag{2}$$

where $B = \frac{(A+A^T)}{2}$, $H = V\Sigma V^*$ and $B = W\Sigma V^*$.

Here, $H$ is the symmetric polar factor of matrix $B$ where the polar decomposition of $B = UH$ $(U^T U = I\ , H = H^T \geq 0)$ . $U$ represents the unitary matrix and $H$ represents the hermitian matrix with PSD property and $B = W\Sigma V^*$ using singular value decomposition (SVD) where $V^* = V^T$ is the conjugate transpose of matrix $V$.

To ensure that the corrected sample covariance matrix is not too far from the actual sample covariance matrix, the difference of the Frobenius norm, $\|A\|_F = \sqrt{\sum_{i,j} a_{ij}^2}$, of the two matrices is reported in the software. Upon ensuring that the covariance matrix satisfies the PSD property, graphical LASSO in "huge" R package is implemented. The function requires a specification of lambda values to tune the $L_1$-penalty term in the model and computes the corresponding penalised log-likelihoods at all lambda values. Model selection criteria, including AIC, BIC, e-BIC and cross-validation (CV), are used to help users to select an optimal regularization parameter $\lambda$. Users may choose to specify their own vector of lambda values, otherwise, the software automatically generates a grid of 30 lambda values that is exponentially decreasing from 1 to 0.01:

$$\lambda_{grid} = \{\lambda_1, \lambda_2, \ldots, \lambda_{30}\} = exp\,\{log(1)\,, log(0.853)\,, \ldots, log(0.01)\} \qquad (3)$$

As a visual aid, a calibration plot is produced as a portable document format (PDF) with all four model selection criteria plotted against the vector of lambda values and users can re-calibrate the lambda values and specify the model selection criterion to use to select the optimal regularization parameter.

After selecting an optimal regularization parameter $\lambda$, the final GLASSO model is fitted and the estimated sparse inverse covariance matrix, $\widehat{\Omega}$, is produced. The non-zero entries in this matrix are then converted into an edge-level network file to be used for the predictive analysis module. In addition, a partial correlation matrix is produced as output to provide a measure of confounding-free correlation for every pair of data features present in the network. For entries where the estimated inverse covariance matrix are shrunken to zero, the partial correlations remain as zero. The partial correlation between data features $i$ and $j$ is calculated as follows:[8, 9]

$$\tilde{\rho}_{i,j} = \frac{-\widehat{\omega}_{i,j}}{\sqrt{\widehat{\omega}_{i,i}\,\widehat{\omega}_{j,j}}} \qquad (4)$$

where $\widehat{\omega}_{i,j}$ represents the $(i,j)^{th}$ entry in the estimated precision matrix, $\widehat{\Omega}$. Then, heat maps illustrating the derivation of the sample covariance matrix to the estimated nearest PD covariance matrix, the estimated precision matrix and the corresponding adjacency matrix, are also produced in PDF files.

***Hybrid approach***

The hybrid approach is a semi-supervised approach where the prior knowledge of the network is combined with the estimated network by the supervised approach above into a single adjacency matrix. First, a network file is required as input as the prior (matrix $\boldsymbol{P}$), then the supervised approach is carried out to obtain an estimated precision matrix (matrix $\widehat{\boldsymbol{\Omega}}$). Let us first define the following matrices:

Prior network: Matrix $\boldsymbol{P}_{p\times p} = (p_{ij}) \in \{-1, 0, 1\}$

Estimated Precision network: Matrix $\widehat{\boldsymbol{\Omega}}_{p\times p} = (\widehat{\omega}_{i,j}) \in [-1, 1]$

Sample covariance matrix: Matrix $\mathbf{S}_{p\times p} = (s_{ij}) \in [-1, 1]$.

Then we can create the following matrices:

$$\text{Matrix } \boldsymbol{A}: (a_{ij}) = \begin{cases} sign(\tilde{\rho}_{i,j}), & if\ i \neq j \\ 0, & if\ i = j, \end{cases}$$

$$\text{Matrix } \boldsymbol{U}: (u_{ij}) = sign(a_{ij} + p_{ij})$$

where the entries in $\boldsymbol{A}$ denote the sign of the partial correlation and the entries in $\boldsymbol{U}$ represent the agreement in the signs of the non-zero entries in matrix $\boldsymbol{A}$ and $\boldsymbol{P}$. If the direction of association between feature *i* and *j* is different in the prior from what is observed in the data (i.e. $a_{ij} = 1, p_{ij} = -1$ or $a_{ij} = -1, p_{ij} = 1$), the entry becomes zero.

We extract the corresponding elementwise sample covariances in $\boldsymbol{S}$ from the non-zero entries in matrix $\boldsymbol{U}$ by defining the Hadamard product of the two matrices to result in a non-negative matrix:

$$\boldsymbol{B} = \boldsymbol{U} \circ \boldsymbol{S} \tag{5}$$

A score matrix $\boldsymbol{S}^*$, constrained between 0 to 1, can be computed as follows:

$$\text{Matrix } \boldsymbol{S}^* = (s_{ij}^*) \begin{cases} -\frac{|b'_{ij}|}{b_{max}} & if\ a_{ij} = 0, p_{ij} \neq 0 \\ \frac{|b'_{ij}|}{b_{max}} & otherwise, \end{cases} \tag{6}$$

where $\boldsymbol{B}' = (b'_{ij}) = \boldsymbol{BWB}^T$ and $b_{\max} = \max_{\boldsymbol{i,j}} \boldsymbol{b}'_{\boldsymbol{ij}}$. Here, the matrix $\boldsymbol{W} = \frac{1}{2}(|\boldsymbol{A} + \boldsymbol{P}|)$ is a weight matrix taking values 0, 0.5 and 1.

Lastly, a fused matrix $\boldsymbol{F}$ is computed by:

$$\boldsymbol{F} = \boldsymbol{U} \circ (\boldsymbol{P} + \boldsymbol{S}^*) \tag{7}$$

adding to the prior if the edge is supported by what is estimated in $\widehat{\Omega}$ (i.e. $\widehat{\omega}_{i,j} \neq 0$), or penalizing the prior if the edge is present in the prior but not supported by the estimated network (i.e. $\widehat{\omega}_{i,j} = 0$).

The final network is generated by forming an edge between feature $i$ and $j$ if the absolute value in $(i,j)^{th}$ entry of matrix $\boldsymbol{F}$ is at least 0.5 and the direction of association of the edge is determined by the sign of the entry. Heatmaps illustrating the derivation of the sample covariance matrix to the estimated precision matrix, the input prior matrix, the product fused matrix and the corresponding adjacency matrix, are produced as a graphical output at the end.

### Model selection criterion

To optimise the regularization parameter, $\lambda$, for the penalised log-likelihood in GLASSO that controls for the sparsity of the precision matrix, several model selection criteria are used, including Akaike Information Criterion (AIC) and Bayesian Information Criterion (BIC) [10, 11]. However, when dealing with molecular data, we often encounter the curse of the high dimensionality where the size of the feature space, *p*, is much greater than the sample size, *n*. Hence, other criteria such as extended Bayesian information criterion (e-BIC) and *K*-fold CV may be more relevant.

***Akaike Information Criterion, AIC***

AIC is a model selection criterion which penalises for overfitting of the model. The formula for AIC is:

$$AIC_\lambda = 2k - 2ln(\mathcal{L}) \quad (8)$$

where $k$ is the number of non-zero elements in the estimated precision matrix ($\widehat{\Omega}$) and $\mathcal{L}$ is the likelihood function without the penalty term. The best regularised parameter would correspond to the model which yield the lowest AIC value.

***Bayesian Information Criterion, BIC***

BIC is similar to AIC but penalises more against over-parameterisation. Due to the heavier penalty, linear regression models chosen by BIC tends to be the same or simpler models than the one chosen by AIC.[12] The formula for BIC is

$$BIC_\lambda = k\,ln(n) - 2ln(\mathcal{L}) \quad (9)$$

where $n$ is the sample size. One underlying assumption of BIC is that the observations should be independent and identically distributed.[13] Otherwise, the effective sample size, $n$, should be adjusted accordingly. Then, the regularised parameter should be picked with the model which has the lowest BIC value.

***Extended Bayesian Information Criterion, e-BIC***

Another recently proposed model selection criterion is the extended BIC, which came about after Foyel *et al.* and Chen *et al.* showed that AIC and BIC lose consistency when the feature space exceeds the sample size (i.e. $p \gg n$). They propose a modification to the BIC formula as follows:[11, 14]

$$e\text{-}BIC_\lambda = k\,ln(n) - 2ln(\mathcal{L}) + 4k\gamma\,log\,(p) \quad (10)$$

where $0 \leq \gamma \leq 1$, with higher values yielding simpler and more parsimonious models. When $\gamma = 0$, the formula reverts to BIC and $\gamma = 0.5$ is suggested to be used for more consistent models.[11] Similar to both AIC and BIC, the best regularised parameter corresponds to the model with the smallest e-BIC value.

***K*-fold Cross-Validation, CV**

An alternative approach to estimate a regularised parameter is through learning directly from the data. In 2009, Fan *et al.*[15] proposed a *K*-fold cross validation score given by:

$$CV(\lambda) = \sum_{k=1}^{K} n_k \, log\left|\hat{\Omega}_{-k}(\lambda)\right| - \sum_{i \in T_k} \left(x^{(i)}\right)^T \hat{\Omega}_{-k}(\lambda) \, x^{(i)} \qquad (11)$$

where $\log|\Omega| = \log(\det(\Omega))$, $n_k$ denotes the sample size and the set $T_k$ represents the test dataset in the $k^{th}$ fold. Here, $\widehat{\Omega}_{-k}(\lambda)$ is the precision matrix estimated by using the training dataset, $\cup_{j \neq k} T_j$. Typically, a five-fold cross-validation is recommended unless the sample size is too small and the regularised parameter is chosen at the value with the largest CV score.

### Software architecture

iOmicsPASSv2plus is developed as an R package and the software is publicly available on GitHub (https://github.com/cssblab/iOmicsPASS). The package includes several modules to guide users to install, create an input parameter file containing the directory to which all the necessary files are and execute the program. The main software iOmicsPASS is still however coded in C++, and the functions enable users to execute the program from the R console, making it more user-friendly for individuals who are less comfortable with the command-line option. Below is the list of the functions included in the R package:

`----- INSTALL.iOmicsPASS() -----`

This function calls system to build and compile the C++ program from the specified directory. In Windows, it creates a Windows executable (.exe) and in Mac OS, it creates a compiled binary in the */bin* directory.

`----- NetDeconvolute() -----`

This function carries out the network inference module by first standardizing the input data (i.e. inputDat) and combining multiple datasets into a single matrix. PCA is then carried out to screen for outlying samples that are more than 4 SDs away from the median of the first 2 PCs. After this, a supervised or hybrid approach is performed to estimate the adjacency matrix from the precision matrix or fused matrix, respectively, and to form the resulting network.

`----- createPrior() -----`

This function fits a logistic model for a binary outcome or a multiple logistic model for a phenotypic group with more than 2 outcomes, adjusting for the user-specified clinical variables. User has to specify which variables are categorical and thus need to be converted to factor variables, otherwise they are treated as continuous variables. It then calculates the probability of every sample's membership to each phenotypic outcome based on the fitted model. The probabilities calculated can be fed into `run.iOmicsPASS()` to be used as prior in the discriminant model for classification.

The data can also be split into a training and test dataset where the logistic model is fitted on the training dataset and the classification probabilities are calculated on the test dataset, by using the option "`predict=TRUE`". The probabilities calculated can be fed into `Predict.iOmicsPASS()` function as prior probabilities used in the discriminant model to assign test samples to the phenotypic outcome groups in the training model.

----- `createInputParam()` -----

This function creates an input parameter file needed to run iOmicsPASS with user's specifications and the directory containing the required input files.

----- `run.iOmicsPASS()` -----

This function calls system and execute the compiled binary, performing predictive analysis to identify network signatures that can separate the phenotypic groups of interest. The software may take some time if the dimensionality of the network is very large. It is advised to first run CV once to create a misclassification error plot by setting `Cross.Validate=TRUE`, then turn off the CV option and re-specify a suitable threshold that strikes a good balance between change in mean CV error and the number of selected edges. Its function also outputs the key model parameters required for prediction module below.

----- `Predict.iOmicsPASS()` -----

This function requires the model parameters associated with the network signatures obtained from a training dataset. Using the same network features in the test data to compute a co-expression matrix, class probabilities are computed to assign each new sample to a phenotypic group.

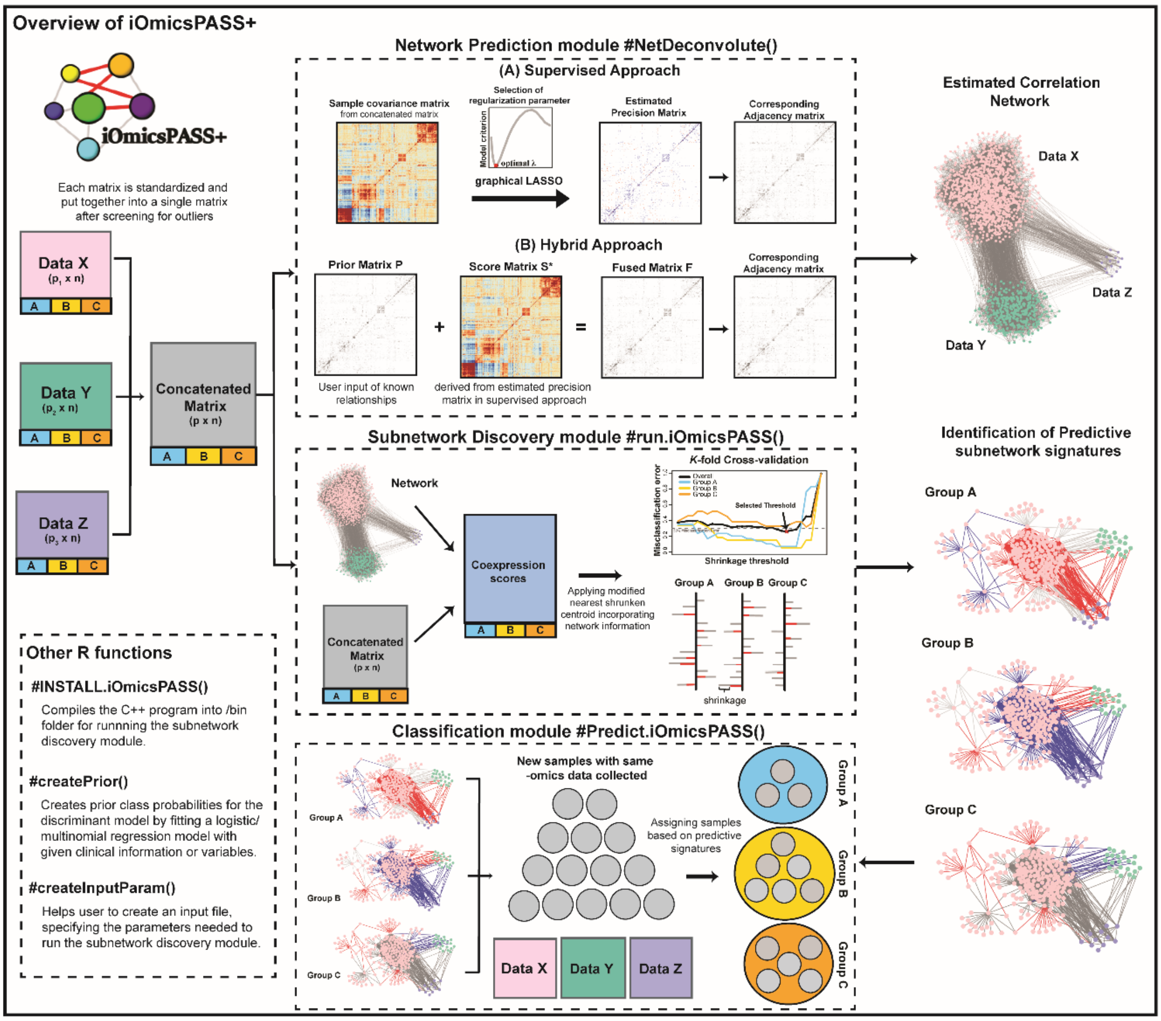
